# Genomic epidemiology of SARS-CoV-2 in the United Arab Emirates reveals novel virus mutation, patterns of co-infection and tissue specific host innate immune response

**DOI:** 10.1101/2021.03.09.21252822

**Authors:** Rong Liu, Pei Wu, Pauline Ogrodzki, Sally Mahmoud, Ke Liang, Pengjuan Liu, Stephen S. Francis, Hanif Khalak, Denghui Liu, Junhua Li, Tao Ma, Fang Chen, Weibin Liu, Xinyu Huang, Wenjun He, Zhaorong Yuan, Nan Qiao, Xin Meng, Budoor Alqarni, Javier Quilez, Vinay Kusuma, Long Lin, Xin Jin, Chongguang Yang, Xavier Anton, Ashish Koshy, Huanming Yang, Xun Xu, Jian Wang, Peng Xiao, Nawal Ahmed Mohamed Al Kaabi, Mohammed Saifuddin Fasihuddin, Francis Amirtharaj Selvaraj, Stefan Weber, Farida Ismail Al Hosani, Siyang Liu, Walid Abbas Zaher

## Abstract

To unravel the source of SARS-CoV-2 introduction and the pattern of its spreading and evolution in the United Arab Emirates, we conducted meta-transcriptome sequencing of 1,067 nasopharyngeal swab samples collected between May 9th and Jun 29th, 2020 during the first peak of the local COVID-19 epidemic. We identified global clade distribution and eleven novel genetic variants that were almost absent in the rest of the world defined five subclades specific to the UAE viral population. Cross-settlement human-to-human transmission was related to the local business activity. Perhaps surprisingly, at least 5% of the population were co-infected by SARS-CoV-2 of multiple clades within the same host. We also discovered an enrichment of cytosine-to-uracil mutation among the viral population collected from the nasopharynx, that is different from the adenosine-to-inosine change previously reported in the bronchoalveolar lavage fluid samples and a previously unidentified upregulation of APOBEC4 expression in nasopharynx among infected patients, indicating the innate immune host response mediated by ADAR and APOBEC gene families could be tissue-specific. The genomic epidemiological and molecular biological knowledge reported here provides new insights for the SARS-CoV-2 evolution and transmission and points out future direction on host-pathogen interaction investigation.

## Introduction

The coronavirus disease 2019 (COVID-19), caused by the infection of severe acute respiratory syndrome coronavirus 2 (SARS-CoV-2)(*1*), has become the largest outbreak since the 1918 Spanish influenza pandemic(*2*). It has resulted in 131.83 million cases and 2.86 million death, as of March, 2021(*3*). Patients infected by SARS-CoV-2 can experience a number of serious respiratory illnesses and have in many cases died from complications related to the infection(*4*). There are no specific therapeutics or fully validated vaccines available for its control to date(*5, 6*). Dynamic transmission modelling considering seasonal variation, immunity and intervention suggests a high possibility of continuing waves of resurgence until the year 2025(*7*).

Genomic epidemiology using massively parallel high-throughput sequencing technologies (MPS) and associated analyses and bioinformatics tools have been used to understand the rapid spread and evolution of the virus at a larger scale than ever before(*8, 9*). Public repositories including GISAID have enabled fast release and sharing of SARS-CoV-2 genome sequences(*10*). Those efforts provide valuable information to researchers and public health officials for global outbreak responses. Nevertheless, there are new questions arising regarding the virus’ ongoing breadth of transmission, its evolution inter- and intra-host, as well as host-pathogen interactions. The genetic diversity of global viral strains is largely underestimated given the lack of real-time sequencing capability in most of the world, resulting in a disproportional under-study of viral populations in under- and recently-developed countries. As a consequence, there is limited information on novel and common genetic variation in those areas where virus rapidly evolves and is subjected to natural selection, as it encounters human hosts with diverse genetic background and an environment with varying temperature and humidity levels(*11, 12*). Most published research since the start of the pandemic has focused on inter-host phylogenetics based on the assumption that only one strain of the virus is present in the sample. Intra-host viral genetic diversity and the prevalence of coinfection has not been established via sufficiently large cohort despite the possibility that it might impact clinical outcomes and potentially enable higher resolution analysis in the who-infects-whom transmission chain(*13*). Finally, while understanding how the host response to the virus will help to combat the disease, innate immune response process such as the host-dependent RNA-editing mechanism has only been investigated among limited sample cases(*14*).

The United Arab Emirates (UAE) is one of the world’s most famous international hubs for business and travel and is the first country to approve a Chinese COVID-19 vaccine. Despite a long-lasting period of epidemic, only a few of the SARS-CoV-2 samples were sequenced and the transmission and evolution patterns of the virus in this area is unknown. The first case of SARS-CoV-2 was detected in the country on January 29^th^, 2020 (**Figure 1**). The subsequent outbreaks infected over sixty thousand individuals by the end of June 2020 and three hundred thousand individuals by the end of December 2020(*3*). Since March 2020, the UAE public health authorities have adopted a series of strict regulations to reduce human-to-human transmission, including airport lockdown and national curfew. On the other hand, due to economic pressures, a few international flights reopened gradually in June 2020, which may be one of the reasons for the subsequent small second peak during June and August. The most outstanding third epidemic peak were observed during the December Christmas time in 2020. There have been 2-4 thousand newly confirmed cases in the country since Christmas. Since the very beginning, as a response to the pandemic, several high-throughput molecular technologies have been adopted in the UAE to extensively monitor the viral spread and for rapid screening of infected patients. A nationwide RT-qPCR screening program conducting ten thousand tests daily was launched on March 31^st^ 2020. Almost simultaneously, a high-throughput sequencing laboratory with 12-18Tbases/day capacity was established in early April 2020, enabling meta-transcriptome sequencing of up to 192 samples in 24 hours.

**Figure 1.**
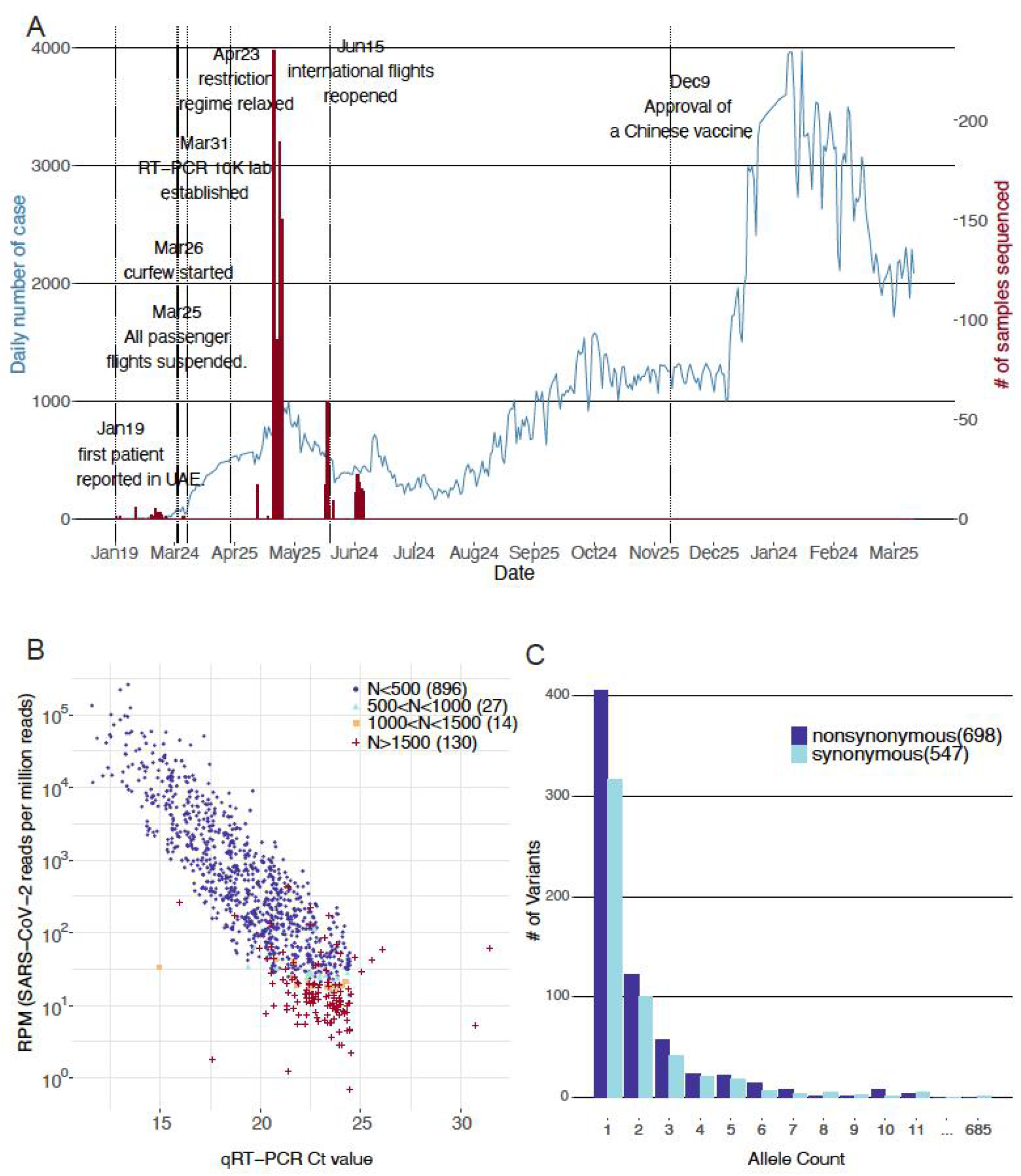
COVID-19 outbreak in the United Arab Emirates and the samples subjected for sequencing in this study. (A) Number of confirmed infected cases in the UAE (N=461,444) until Mar 31st 2021 was shown in the blue line and the number of subjects sequenced by meta-transcriptomic sequencing (N=1,067) was shown in the red bars. Important dates reflecting governmental responses were marked in black text. (B) Assembly quality of the 1,067 viral genomes as a function of the RT-PCR Ct value and SARS-CoV-2 reads per million sequencing reads. Color represents assembly quality stratified by the number of gaps. (C) Allele frequency spectrum of the 1,245 genetic variants identified from the 896 assemblies with less than 2% gaps.

To understand the transmission and infection dynamics of SARS-CoV-2 within the UAE and in relation to other countries, during April and July, 2020, we randomly collected 1,067 nasopharyngeal specimens from SARS-CoV-2 positive patients from the RT-qPCR screening program and conducted meta-transcriptomic sequencing. Our main scientific questions include (1) What is the virus genetic diversity and transmission pattern in the UAE during the first peak of the epidemic (2) What is the extent of co-infection of multiple SARS-CoV-2 variants in this international travel hub (3) Is there any innate immune host response to the SARS-CoV-2 infection that can be detected using the meta-transcriptomic sequencing, which contains both the host and the viral gene expression information.

## Results

### Assembly and variant detection of SARS-CoV-2 genome from deep meta-transcriptome sequencing of 1,067 nasopharyngeal swab samples

A total of 1,067 nasopharyngeal swab samples collected from SARS-CoV-2 positive patients between May 7^th^ and June 29^th^ 2020 in Abu Dhabi were sequenced (**Figure 1A)**. Their sequencing quality metrics were summarized in **Figure S1 and Table S1**. We obtained high quality assemblies (gap proportion < 2%) for the majority of the samples (n= 896, 84.0%). In brief, using the 29891nt SARS-CoV-2 reference genome (IVDC-HB-01), we have successfully assembled all 1,067 SARS-CoV-2 consensus genomes as follows-896 assemblies with gaps less than 500nt (gap proportion < 2%), 27 assemblies with gap less than 1000nt (gap proportion < 4%), 14 assemblies with gaps less than 1500nt (gap proportion ∼5%) and 130 assemblies with gaps greater than 1500nt (**Figure 1B**). As expected, quality of the genome assemblies was closely related to the sample viral load as measured by reads per million (RPM) and qRT-PCR Ct values (**Figure 1B, Figure S2**). A set of 3 samples (id:0555, 0919 and 0945) showed low viral loads (Ct<19) with unexpectedly poor assemblies (gaps>1500nt), likely due to RNA degradation as many of the sequenced reads were filtered out due to low complexity, i.e. high polyA proportion (**Table S1**).

The distribution of gaps identified in the sequences indicates low sequencing coverage over the 5’ and the 3’ ends of the genomes, which was found to be a common occurrence in all world-wide assemblies reported in GISAID. We also notice a significantly higher number of gaps around the 20,000nt position for 27.1% of the assemblies submitted to GISAID, which were not observed in our assemblies (**Figure S3**). Among the selected 896 assemblies with the highest quality (gap proportion < 2%), we identified a total of 1,245 genetic variants consisting of 698 non-synonymous and 547 synonymous variants when compared to the SARS-CoV-2 reference genome (IVDC-HB-01), (**Figure 1C, Table S2**). The number of variants per sample ranged from 1 to 24 with a median number of 11 (**Figure S4**). Very few genomes carried non-single nucleotide variants. There was one 2nt insertion in one sample 1069 and six deletions identified in fourteen samples 0188,0236,0252, 0290, 0305, 0339, 0512, 0536, 0757, 0758, 0761, 0763, 0785 and 1092, the largest being a 4nt deletion present in seven of the fourteen samples (**Figure S5**). The consensus variants identified from the technical replicates were exactly the same (**Table S3**), and given a 4% alternative allele frequency threshold, the concordance rate of intra-host genetic variant detection reaches 100% (**Figure S6**). The number of variants that we identified per sample did not correlate with the sequencing depth (squared pearson correlation coefficient R^2^∼0.02) (**Figure S7**).

### Global clade composition and five novel subclades associated with eleven novel common genetic variants in the UAE SARS-CoV-2 population

Likely due to fast population expansion with a short period, we discovered that 395 out of the 896 genomes (44.1%) assembled in our study shared an identical genome sequence with at least one other assembled genome (**Table S4**). For the purpose of downstream phylogenetic analysis, we filtered the 896 genome sequences as to keep only unique sequences, resulting in 637 unique genome sequences. We constructed a maximum likelihood phylogenetic tree including, 1) the 637 SARS-CoV-2 unique genomes and collected assembled in our study between May 7^th^ and June 29^th^ 2020 in Abu Dhabi, 2) the 52 nearest relative world-wide genomes identified from GISAID between February 2^nd^ and April 24^th^ 2020 (**Table S6, Figure S8**), and 3) 25 genomes collected from the nearby Dubai Emirate between January 29^th^ and March 18^th^ 2020 (*15*). We identified the five dominant clades worldwide (*16, 17*) in the UAE viral population sequenced in this study **(Figure 2A)**. A total of 13 (2.04%) and 140 viral genomes (21.98%) out of the 637 genomes were clustered as clade 19A and clade 19B, respectively, the two earliest clades first reported in China, Asia(*18*), while the rest of the genomes sequences were classified in the clades 20A (N=52, 8.16%), 20B (N=428, 67.19%) and 20C (N=4, 0.63%), which were first reported and became prevalent in Europe and North America^4,16^. Three samples in clade 19A, i.e. samples 0134, 0135 and 0565, harbored a higher number of mutations; 20, 19 and 19, respectively, compared to the calculated average of 11 variants per genome. The closest strain found to these three samples was SARS-CoV-2 USA/WA-S771/2020 reported in Washington, DC, United States on April 13^th^, 2020 (**Table S6**). The high level of mutations occurring in these samples compared to the rest of the UAE genomes, indicates a different introduction of strains within the same clade.

**Figure 2.**
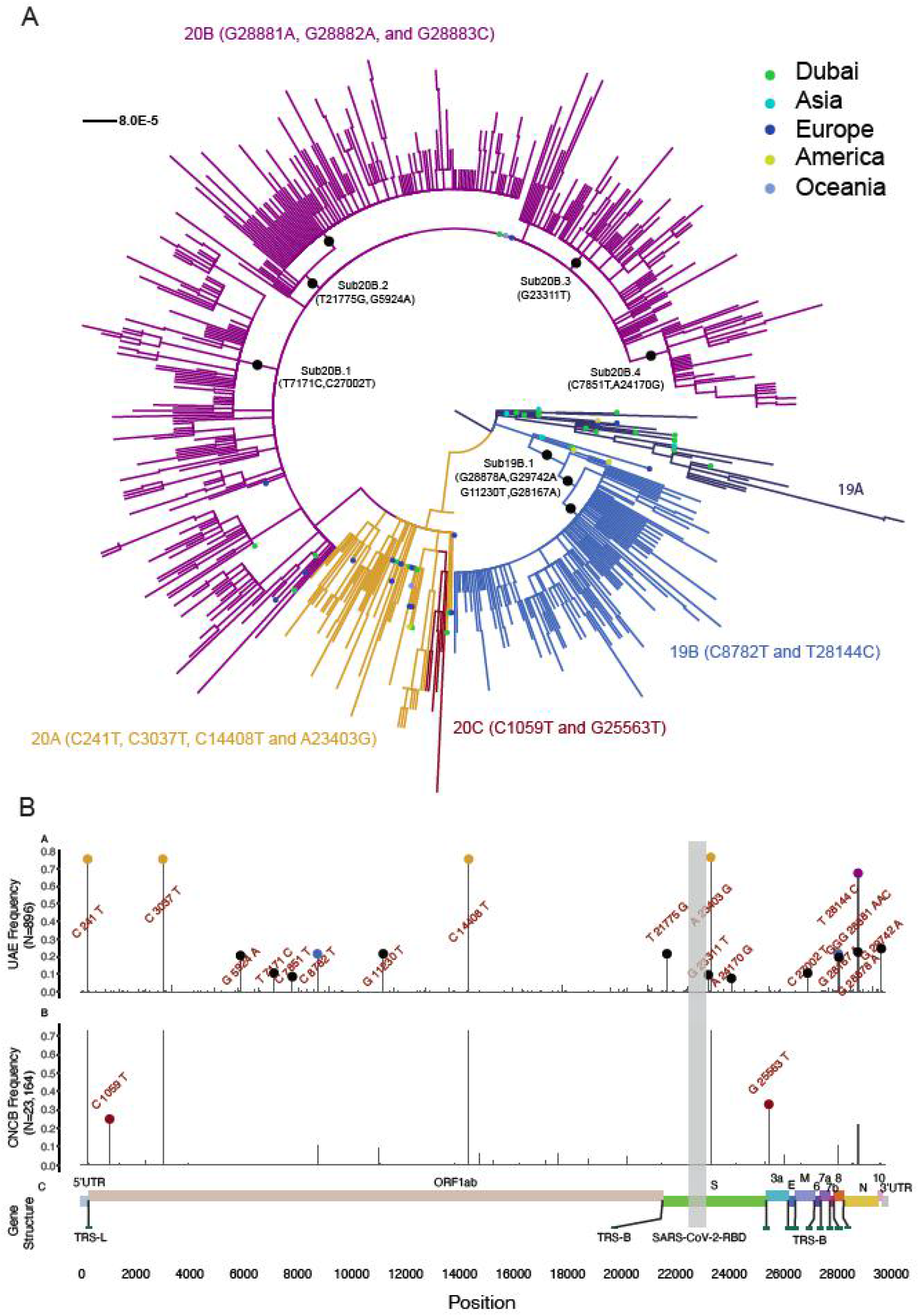
Phylogenetic analysis of the sequenced UAE viral population during May and June. (A). Maximum likelihood tree of the 637 unique viral genomes with less than 2% gaps and 52 closest relatives from GISAID. Each line indicates a sample colored by the five dominant viral clades worldwide, annotated with the clade definitive genetic variation. The subclade-definitive genetic variations were also marked in black. The closest relatives from GISAID were marked by a dot colored by geographical district reported for the viral sample. (B). Comparison of the alternative allele frequency of the 1,245 viral genetic variants between the 896 high quality UAE viral genomes and the 23,164 viral genomes from the globe downloaded from the China National Center for Bioinformation. Nomenclature of the clades was detailed in Supplementary Notes.

There were five large sub-clades involving more than half of the collected samples (381 out of the 637 unique viral genomes, 59.81%) (**Figure 2A)**, differentiated by eleven mutations that were common in the UAE viral population (allele frequency > 5%) and that were significantly less common among the worldwide viral population (P < 3.94e-82, Fisher exact test) (**Figure 2B, Table 1**). The five sub-clades were (1) 19B.1 which consisted of 17.27% of the 637 UAE unique samples, harboring the G28878A, G29742A, G11230T and G28167A mutations; (2) 20B.1 which consisted of 8.48% of the samples, harboring the T7171C and C27002T mutations; (3) 20B.2 which consisted of 19.15% of the samples, harboring the T21775G and G5924A mutations; (4) 20B.3 which consisted of 8.95% of the samples, harboring the G23311T mutation and (5) 20B.4 which consisted of 5.97% of the samples, harboring the C7851T and the A24170G mutations.

Fortunately, individuals classified as carrying certain subclades of the virus did not display significantly different viral loads in their samples as reflected by the RT-qPCR Ct values (**Figure 3**). These 11 variants that defined the subclades tend to occur in highly conserved regions within the SARS-CoV-2 genome (**Figure S9**). Molecular dynamic analysis of two of the missense variants in the spike protein did not suggest substantially different change of the protein structure between the mutant and the wildtype (**Figure S10, Table S7**). Likely due to a recent occurrence, the temporal change of the mutation allele frequency for the subclade-definitive variants is smaller compared to the clade-definitive variants (**Figure S11-S12**).

**Figure 3.**
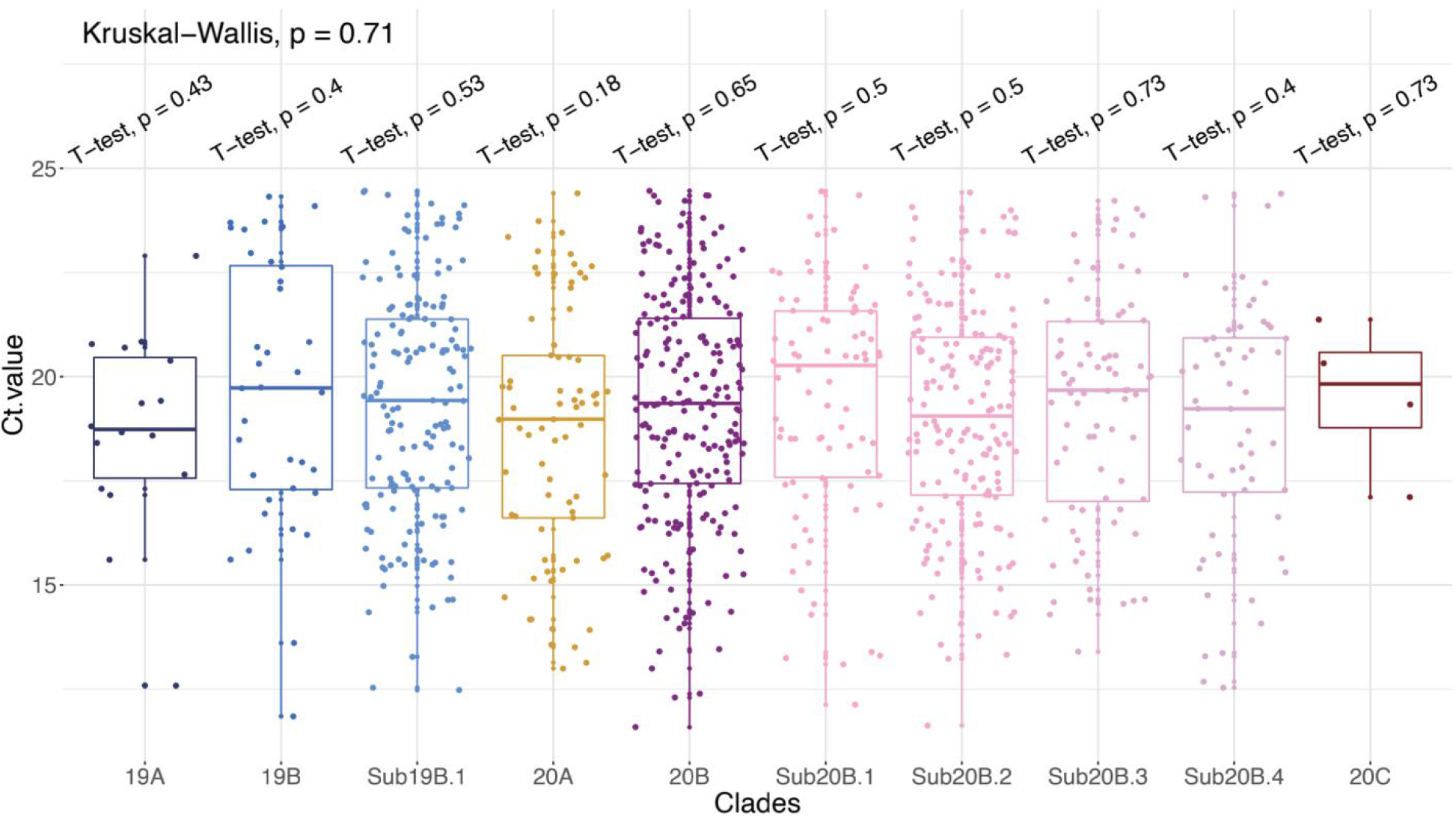
Functional analysis of the unique variants and subclade in the UAE samples. RT-qPCR Ct value distribution for samples in each of the five dominant clades and five subclades. Shown is the p-value using Kruskai-Wallis test and p-value by performing T-test comparing the Ct value for patients carrying certain clade or subclade virus strains with the rest of the patients who didn’t carry the virus belong to a specific clade or subclade.

### Cross-settlement human-to-human transmission contributes to the UAE epidemic

We further investigated human-to-human transmission across 14 settlements from three regions in the Abu Dhabi Emirate and 1 settlement in the Dubai Emirate by constructing the transmission network for 120 samples with geographical and sampling date information (**Figure 4A**). The constructed transmission network indicates prevalent cross-settlement human-to-human transmissions contributing to the epidemic, as within each clade or sub-clade, samples from multiple geographical areas were observed (**Figure 4B**). We also determined the genetic distance using the L1-norm metric that utilized intra-host genetic variation rather than merely the consensus genetic variation, among longitudinal samples (n=24) defined as, same individuals (n=7) sampled multiple times (avg=5.2) over a determined period of time (avg= 4.06 days), and among samples from the same and varying settlements (**Figure 4C**). The median L1-norm genetic distance was smallest among the 24 samples within the longitudinal sampling period, suggesting high levels of stability in viral composition within the same host. As expected, most samples within the same settlement had a genetic distance smaller than the cross-area distance with only two exceptions - samples from the Ghayathi settlement in the AI-Dhafra region and samples from Khabisi in the Dubai emirate, that displayed the largest genetic distance. This is consistent with the fact that those two settlements were relatively less populous compared to the settlements in the Abu Dhabi and AI-Ain regions. The spectrum and the scale of the L1-norm genetic distance is much larger than the genetic computed from the consensus genetic variants although the haplotype information is missing. Due to the small scale of sampling, we didn’t further resolve the transmission network to a finer scale.

**Figure 4.**
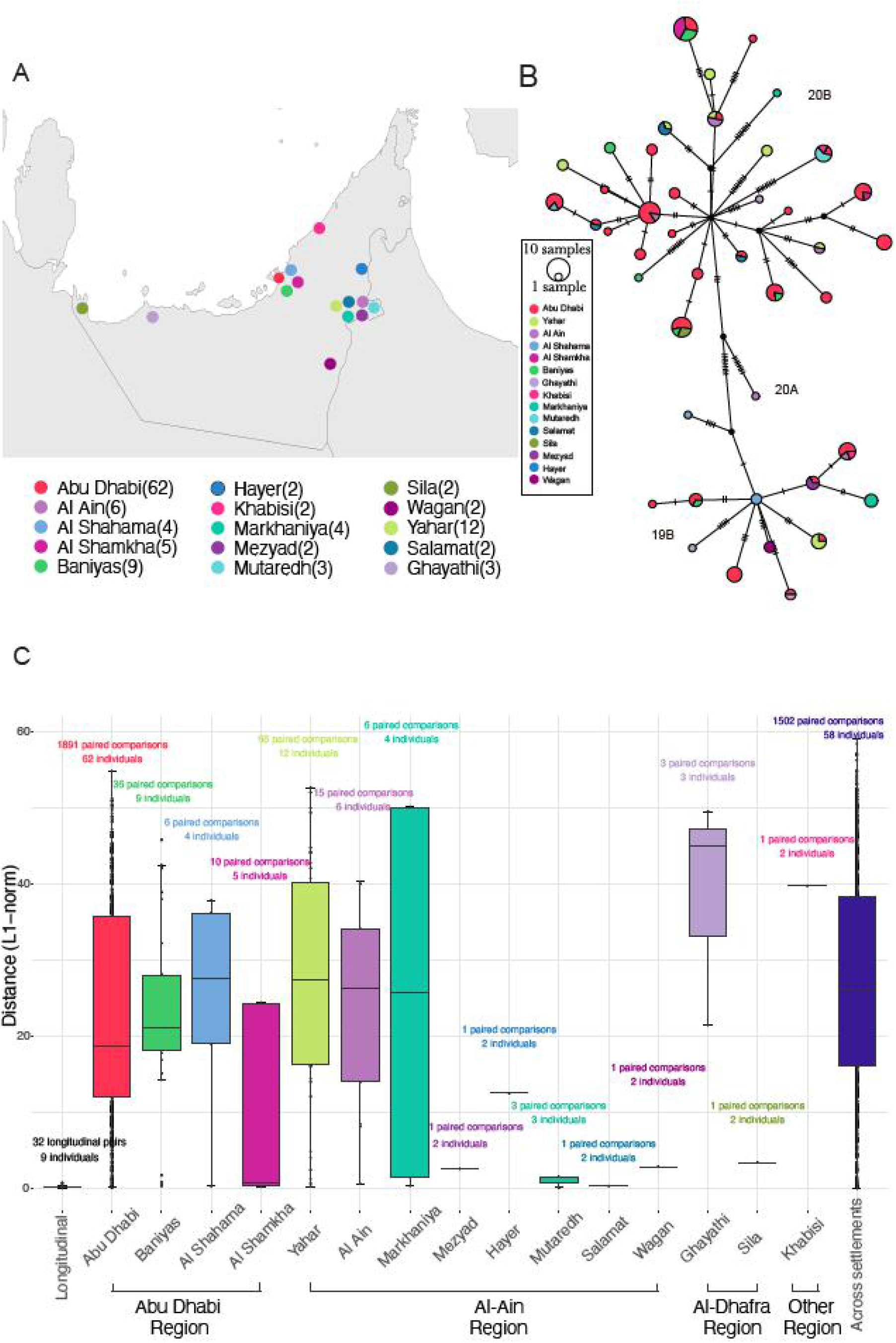
Human-to-human transmission across settlements. (A). Geographical distribution of 120 viral samples with settlement level information in the Abu Dhabi city. (B). Transmission network of the 120 samples colored by settlements. (C). L1-norm genetic distance for longitudinal samples, samples from the same settlements, and samples from different settlements. Among the 130 samples that report settlement level geographical location in Table S5, 10 samples were not displayed because only one sample were collected from that settlement.

### Prevalent co-infection by multiple SARS-CoV-2 variants in the same host

The international hub status of the UAE provides a good opportunity to study the prevalence of multiple SARS-CoV-2 variant co-infection within the same host. We have identified a total of 1,268 intra-host single nucleotide variation (iSNV, with minor allele count of 4 and minor allele frequency greater than 5%) present in 625 out of the 896 samples, ranging from 1 to 26 iSNV per individual with an average of one per individual (**Figure S13**). Although the technical replicates indicate 100% concordance of the iSNV detection at the above threshold, we chose a conservative way of evaluating the prevalence of multiple infection present in the sampled viral population by restricting the definition of co-infection by the co-occurrence of two clades including 19A, 19B, 20A, 20B and 20C (classified using the eleven clade-definitive variants in Figure 2) or subclades (classified using the other eleven sub-clade definitive variants) in the same sample. We found that a total of 48 samples out of the 896 (5%) carried viral variants from more than two distinct clades or subclades (**Figure 5**). The high linkage disequilibrium of the genetic variants that belong to a specific clade indicates the likely presence of a viral variant rather than spontaneous *de novo* mutations. Notably, two of the samples (id: 0855 and 0796) with identical consensus sequence displayed different patterns of multiple infection. Sample 0796 harbored viral genetic variants from clades 19A, 20A, 20B while 0855 harbored variants from clades 20A, and 20B and not from 19A. Samples in the same clade classified by the consensus variants also demonstrate a different pattern of co-infection. For example, for samples in clade 19B, two clusters were observed. One consists of seven samples with multiple infections from several clades (19A, 19B, 20A, 20B) and the other cluster consists of ten samples co-infected with 19B and 20A. For the most prevalent clade 20B viral sub-population, samples could be co-infected by 19A or 20C. Those patterns in Figure 5A largely maintain when using a 0.5% minor allele frequency threshold and the same 4 minor allele support (**Figure S14-S15**), showing a tremendous amount of intra-host genetic diversity underlying the consensus genomes of the host.

**Figure 5.**
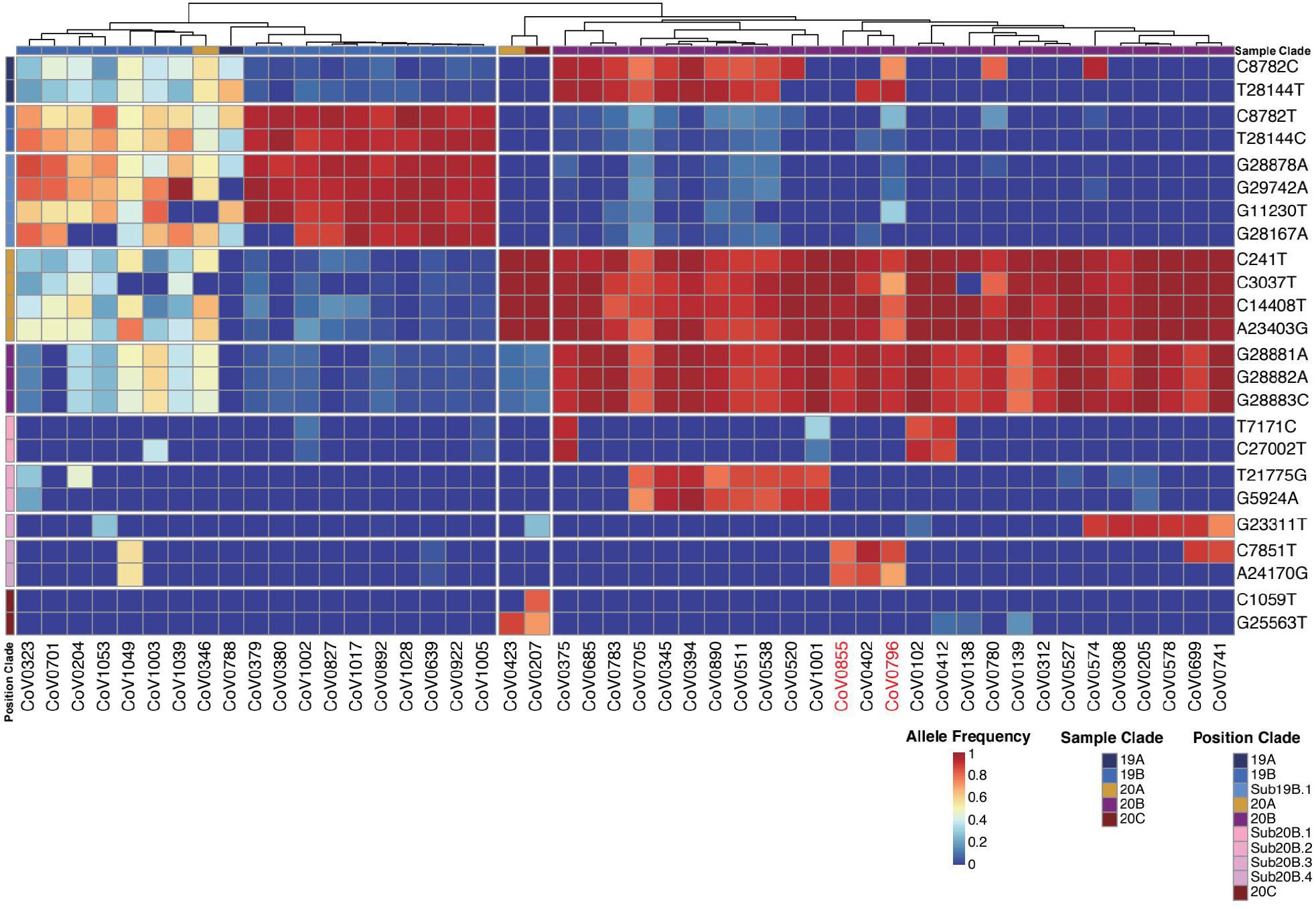
Co-infection with multiple SARS-CoV-2 variants. Evidence for human-to-human transmission of multiple SARS-CoV-2 variants were established using the clade and sub-clade definitive viral genetic variants. Columns display the de-identified sample ID that carried more than one SARS-CoV-2 viral variants in the nasopharyngeal swab sampling (N=48). Color bar shows the viral clade assigned to the individual, according to the consensus viral sequence, reflecting the dominant clade in one sample. Rows indicate the eleven clade-definitive and eleven sub-clade definitive variants. Heatmap color, ranging from red to blue, suggests the allelic proportion of the derived allele of the iSNV. The ID of two longitudinal samples were marked in red.

### The innate immune host response to SARS-CoV-2 infection may be tissue-specific and associated with the upregulated gene expression of *APOBEC4*

We further investigated detectable innate immune host response to SARS-CoV-2 infection utilizing information that can be extracted from the meta-transcriptomic sequencing. A recent publication by Giorgio *et al*. reported evidence of RNA editing in bronchoalveolar lavage fluid (BALF) from eight patients diagnosed with SARS-CoV-2 infection in Wuhan city, China(*19*). For seven out of the eight samples, they identified a bias of the mutation towards transition, mainly A>G/T>C changes followed by C>T/G>A changes, indicating a deamination effect introduced by ADARs and APOBECs, respectively (**WH BALF in Figure 6A**). In the nasopharyngeal swab sampling of 896 patients in our study, on the contrary, we identified the C>T/G>A as the predominant SNV type that were more likely to be mediated by APOBEC gene family rather than the A>G/T>C effects mediated by the ADARs (**UAE in Figure 6A**). This held true when only mutations that occurred in more than two patients were considered. As expected, the C-to-U changes are biased toward the positive strand, i.e. more C-to-U was observed compared to G-to-A, as APOBECs are supposed to target single stranded RNA(*20*). The observation of a dominant C-to-U changes were replicated in the nasopharyngeal swab samples collected in Spain, Virginia and Ruijin hospitals in Shanghai city, China and the 23,164 high quality sequences collected in GISAID (Supplementary notes), which consistently displayed an enrichment in the C>T/G>A mutations, same as the pattern in the UAE nasal swab samples but different from the Chinese BALF results reported by Giorgio *et al (****Figure 6A***). Additional evidence can be obtained with the observation of cytosine depletion in viral sequences during the past ten months, reflected by an increasing of T and A bases and a decreasing of G and C bases (**Figure S16**).

**Figure 6.**
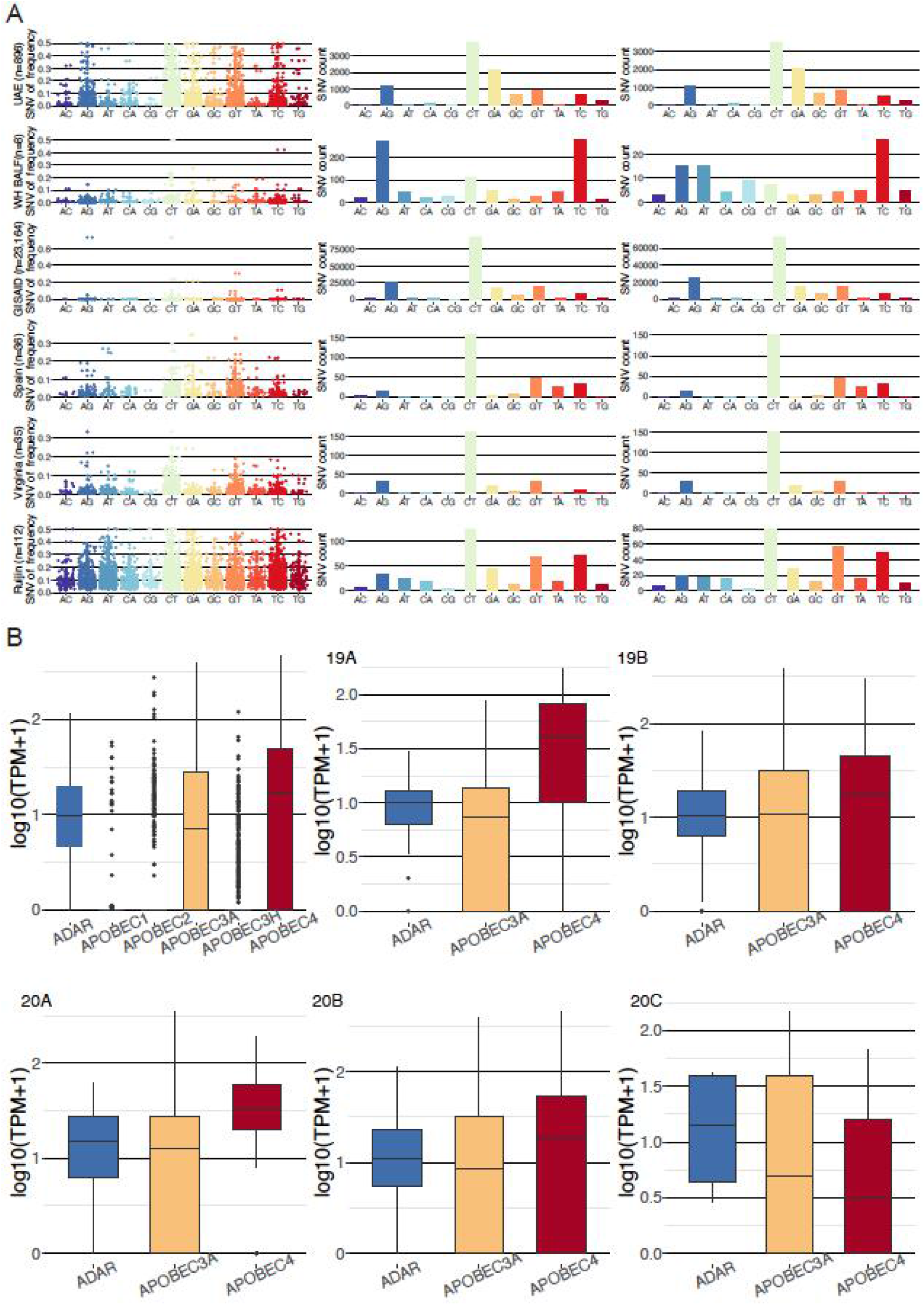
Human innate immune response to SARS-CoV-2 mediated by the ADAR and APOBEC gene families. (A). Allelic faction (Column 1), the number of mutations (Column 2) and the number of recurrent mutations (Column 3) for ten mutation types for six studies arranged by row. UAE: 896 nasal swab samples collected in our study; GISAID: 23,164 viral sequences collected; Spain: 36 nasal swab samples collected in Spain; Virginia: 35 nasal swab samples collected in Virginia and 112 nasal swab samples collected in Ruijin hospital in Shanghai city, China. (B). Host *ADAR* and *APOBEC* gene expression (logarithm of transcript per million) in the nasal swab samples for all and for each of the five clades.

We further investigated if the different patterns observed could be due to the differential gene expression of the *APOBEC* gene families and *ADAR* in the nasopharyngeal swab vs. BALF using public multi-tissue gene expression information from GTEx repository(*21*) and by analyzing the gene expression of *APOBEC* and *ADAR* genes in our sequencing data. According to the GTEx gene expression data among 49 tissues and cells, *ADAR* demonstrated the highest gene expression compared to *APOBEC* gene family in the lung and in the minor salivary gland, the two most relevant tissue compared to the nasopharynx used in our study (**Figure S17**). The GTEx information cannot directly explain the different mutation pattern between the BALF and the nasal swab samples.

Distinct from the GTEx profile obtained from the uninfected individuals (Figure S17), *APOBEC4*(*A4*) displayed the highest average gene expression in the nasal swab samples collected in our study, followed by *ADAR* and *APOBEC3A*, while there were very few samples expressed *APOBEC1, APOBEC2* and *APOBEC3H* (**Figure 6B**). The difference of gene expression is significant between *A4* and the *ADAR* (Wilcoxon test P=7.7e-05) and the largest difference was observed among the individuals carrying clade 20A variants followed by the clade 19B variants (**Figure 6B, Table S8**). In GTEx, *A4* is expressed most prominently in testis, lowly expressed in lung and infrequently expressed in other tissues (**Figure S17**).

The significantly up-regulated *A4* gene expression in the nasopharynx could have been triggered by the SARS-CoV-2 infection. A4 was an under-studied putative cytidine-to-uridine editing enzyme, which cytidine deaminase activity was not as well-known as the APOBEC3A(*22*). The sequencing data not aligned to the SARS-CoV-2 were filtered out from the BALF samples and therefore, we were not able to investigate the gene expression of those host genes in this tissue. That the A4 was previously reported to enhance the replication of HIV-1 indicates its involvement against the RNA virus infection. The high expression of *A4* in nasopharynx may provide the first evidence that the enzyme may be involved as part of the host responses upon the SARS-CoV-2 infection and further experimental analysis is worthwhile to understand its exact functions.

## Discussion

Our analysis of the 1,067 viral genomes collected in the UAE suggest that, during the first quarter of 2020, there were multiple and likely independent introductions of SARS-COV-2. The five dominant global clades of SARS-CoV-2 were all commonly present in the sampled individuals (Figure 2). The highest prevalence of the European dominant clade 20B, followed by the East Asian dominant clade 19B, indicates effects of either a larger founder population size or positive selection. There was substantial local transmission within and between areas in the Abu Dhabi emirate (Figure 4). We have identified 5 new sub-clades, namely; 19B.1, 20B.1, 20B.2, 20B.3 and 20B.4, defined by 11 variants uniquely found within the UAE. Those variants are potentially neutral given that no significantly different viral loads (reflected by the RT-qPCR test) were detected between patients carrying the subclades and those did not (Figure 3).

While consensus sequences tend to be highly similar, intra-host variation adds information which is a promising novel direction for resolving finer-scale transmission networks and studying co-infection of the patients. This study offers the first insight into the prevalence of co-infections of multiple SARS-CoV-2 strains in a large cohort. We observed that at least 5% of the patients were infected by more than one SARS-CoV-2 strain. Within-host co-infection of SARS-CoV-2 variants has been reported in very few studies and with limited sample size. The environment created by the UAE’s “ international hub” status also enables a reliable approach to study co-infection within an individual by different strains of SARS-CoV-2 using clade and sub-clade definitive genetic variants. This raises the importance of carefully collecting valuable epidemiological data worldwide, on the origin and clinical relevance of the multiple infections, and the possibility of further granularity when studying transmission dynamics by utilizing information from multiple strains.

While this study showed that SARS-CoV-2 successfully mutated in the two-month period collection in the United Arab Emirates, it is clear that a large number of mutational changes have taken place in the past 10 months of this pandemic. This would likely result in an immunologic battle between host response and changes in the viral genome potentially leading to important structural changes. We observed a significant accumulation of C-to-U mutations in the nasopharyngeal swab samples collected in this study compared to the early stages of sampling around the globe. This pattern is different to what has been reported in a recent study where an enrichment of A-to-G was followed by T-to-C mutations in seven out of eight BALF samples from Wuhan(*19*). We suspect that tissue-specific gene expression of ADAR and member of the APOBEC protein family may contribute to this observation and discovered that *APOBEC4* was highly expressed in the nasopharynx. Given that APOBEC4 was previously reported to enhance RNA virus replication and was mainly expressed in Testis in an ordinary status, it will be interesting and worthwhile to understand more about its exact function towards the SARS-CoV-2 infection using experimental analysis.

The genomic epidemiological insights from our study will provide a strong basis for the surveillance of emerging mutations within the local viral population. Following the gradual reopening of borders and worldwide travels, the continuous sequencing and identification of allele frequency changes of those variants and additional experimental validation are necessary to verify their biological impacts. Future efforts will be aimed at speeding up the process in providing near real-time molecular surveillance and in the coordination of epidemiological and genomic data to rapidly adapt to SARS-CoV-2 evolution to ensure public safety, adequate diagnosis and accurate pharmaceutical development.

## Methods

### Study design and population

Patients with positive RT-qPCR SARS-COV-2 diagnosis are referred to local designated hospitals administered by the Abu Dhabi Health Services Co (SEHA) and the Department of Health in Abu Dhabi (DOH) for quarantine and treatment. Through a routine surveillance system, all cases of SARS-CoV-2 are reported to the DOH.

In this population-based retrospective study, we have randomly selected 1,067 patients testing positive for SARS-CoV-2 during the months of May and June 2020, regardless of their clinical symptoms. We collected the nasopharyngeal swab samples of the patients from the population screening program and sent them to G42 Biogenix laboratory for RNA extraction using the MGIEasy Magnetic Beads Virus DNA/RNA Extraction Kit (MGI, Shenzhen, China) on MGISP-960 (MGI, Shenzhen, China).

Real-time quantitative PCR (RT-qPCR) was used to quantify viral abundance in the sample, determined by Ct values. The electronic epidemiological meta-data was provided by the DOH using the case report form. The study was approved by the Abu Dhabi COVID19 Research IRB Committee (approval number DOH/CVDC/2020/1945). All analyses were performed on the G42 Health AI computational platform (https://www.g42health.ai/) under local data security and privacy regulations.

### Classification of the SARS-CoV-2 reads from the meta-transcriptome sequencing

Classification, *de novo* assembly and consensus variation detection of the SARS-CoV-2 generally follow the protocol in our previous study^15^. Briefly, total reads were processed using Kraken v0.10.5 (default parameters) with a self-built database of Coronaviridae genomes (including SARS, MERS, and SARS-CoV-2 genome sequences downloaded from GISAID, NCBI, and CNGB) to identify Coronaviridae-like reads in a sensitive manner. Fastp v0.19.5 (parameters: -q 20 -u 20 -n 1 -l 50) and SOAPnuke v1.5.6 (parameters: -l 20 -q 0.2 -E 50 -n 0.02 −5 0 -Q 2 -G -d) were used to remove low-quality reads, duplications, and adaptor contaminations. Low-complexity reads were then removed using PRINSEQ v0.20.4 (parameters: -lc_method dust -lc_threshold 7).

### Alignment to reference genome

Reads aligned to SARS-CoV-2 reference genome (BetaCoV/Wuhan/IVDC-HB-01/2019|EPI_ISL_402119) were classified as SARS-CoV-2 reads. Sequencing depth was measured using samtools depth using the default parameters. Samples that exhibited 10-fold average sequencing depth after filtration were accepted for downstream analyses. Reads per million (RPM) belonging to the SARS-CoV-2 was estimated by dividing the reads aligned to SARS-CoV-2 by the total number of reads generated from the same sample.

### Genome assembly

The BetaCoV/Wuhan/IVDC-HB-01/2019|EPI_ISL_402119 sequence was used as the virus reference genome. The IVDC-HB-01 reference lacks 12 A nucleotides at the end compared to Wuhan/Hu-1/2019 and consists of 24 more sequences at the 5’ beginning compared to Wuhan/WH01/2019. SARS-CoV-2 consensus sequences were generated using Pilon v1.23 (parameters: --changes –vcf --changes --vcf --mindepth 10 --fix all, amb)^16^. Nucleotide positions with sequencing depth < 10× were masked as ambiguous base N. We have also applied *de novo* assembly of the Coronaviridae-like reads from samples with < 100× average sequencing depth using SPAdes (v3.14.0) with the default settings. The Coronaviridae-like reads of samples with > 100× average sequencing depth across SARS-CoV-2 genome were subsampled to achieve 100× sequencing depth before being assembled. However, the assembled genomes are enriched of errors and therefore we didn’t use those assembled sequences in the downstream analysis.

### Consensus variation detection and annotation

Pilon generates a variant calling formatted file for recording the consensus variation. To verify the correctness of those consensus variation calls, we also applied freebayes (v1.3.1) (parameters: -p 1 -q 20 -m 60 --min-coverage 10 -V) to detect genetic variation from the bam file. The low-confidence variants were removed with snippy-vcf_filter (v3.2) (parameters: --minqual 100 --mincov 10 --minfrac 0.8). The correctness of those results was evaluated using the two technical replicates (**Table S3**). The remaining variants in VCF files generated by freebayes were annotated in SARS-CoV-2 genome assemblies and consensus sequences with SNVeff (v4.3) using default parameters^17^. Jalview (v1.8.3) was used to perform multiple sequence alignment and estimate the conservativeness score of the mutations^18^.

### Intra-host variation detection

We applied reditools^19^ to compute the sequencing depth of the four A, C, G, T bases (parameters: python2.7 reditools.py -f sample.bam -o sample.count.txt -S -s 0 -os 4 -r ref.fa -q 25 -bq 35 -mbp 15 -Mbp 15). The intra-host genetic variation was detected using reditools(*24*) with a minimum frequency of 5% and 4 copies of minor alleles. We have applied three technical replicates for two samples to evaluate the accuracy of the assembled sequence, the consensus and intra-host genetic variants. This conservative cutoff was decided based on the two sets of technical replicates with examination of concordance (SNV found in both samples) and discordance (SNV found in only one of the two samples) for different frequency thresholds.

### L1-norm genetic distance

We calculate the L1 norm genetic distance by comparing each variant nucleotide position of two samples.

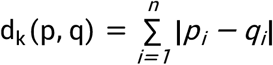

We define dk as the distance measured at position k for comparing samples p and q, and n is the total number of possible nucleotide configurations (A, C, G, T) to calculate the difference in frequency of the same nucleotide in different two samples. For each pair of samples, we use D to represent the sum of the degree of difference in all positions, and N is the sum of the number of variant nucleotides in the two samples.

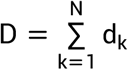

This single number D quantifies the degree of difference in all nucleotide variants between the two samples. We repeated this process for all samples.

### Analysis of host ADAR and APOBEC gene expression

Reads were aligned to the human genome reference (GRCh38) using hisat2 (parameters: --phred64 --no-discordant --no-mixed -I 1 -X 1000 -p 4). Reads aligned to the exons defined by UCSC (gencode.v29.annotation.gtf) were counted (parameters: -s no -f bam -t exon -m union -r name -i gene_id). TPM was defined by the following formula where

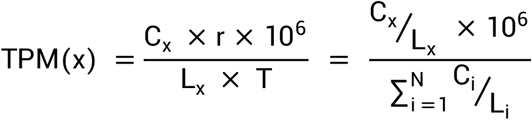

where x refers to a gene or a transcript. R refers to the read length, Cx indicates the number of read pairs aligned to the exons of the gene x. T indicates the length of the gene (kb) divided by the total length of all the genes (kb). Lx indicates the length of gene x.

### Phylogenetic analysis and cross-area transmission inference

From the total 896 assembled high-quality genomes (<2% gap proportion), 637 were unique, therefore considered as different strains, and were used for further phylogenetic analysis. These were aligned to 46,917 genome sequences collected outside of the UAE between January 10^th^ and June 16^th^ 2020 and deposited to the GISAID EpiCoV database (https://www.epicov.org/).

As subset of genome sequences were selected for phylogenetic tree building, including the 637 strains sequenced in this study, the 52 most closely related genome sequences from the alignment analysis against the global 46,917 sequences, and 25 genome sequences also obtained from GISAID that were collected and sequenced in Dubai, UAE, from January 29^th^ to March 15^th^ 2020. We built a maximum likelihood phylogenetic tree using the Nextstrain pipeline; Augur v6.4.3 and MAFFT v7.455 for multiple sequence alignment and IQtree v1.6.12 for phylogenetic tree construction (*25*). FigTree v1.4.4 was used to visualize and annotate the phylogenetic tree. Clades were defined following the Nextstrain nomenclature(*16*). Subclades were further defined in this study based on common variants (>5%) in the UAE but is significantly rarely present in the rest of the world (fisher exact p-value < 4e-82).

Samples with corresponding epidemiological data including patients’ addresses and date of first sample collection were also used to generate median-joining networks for each clades and subclades using PopART (Population Analysis with Reticulate Trees) v1.7. L1-norm genetic distance was computed using the formula previously defined in the influenza study by Poon, *et al* (2016)(*13*), reflecting the sum of the degree of difference for each variant nucleotide position of any two samples.

### Statistical analysis

Fisher exact tests were applied to the 637 unique genomes identified in this study and to 23,164 SARS-CoV-2 genomes collected worldwide from GISAID and curated in the China National Center for Bioinformation (CNCB)(*26*). The tests were used to identify variants that display substantial allele frequency differences between the two sets of genomes sequences; UAE vs. rest of the world. Kruskai-Wallis test was used to compare the RT-qPCR Ct values between clades and subclades.

The distribution of the 10 types of genetic mutations (e.g. A>C, C>G mutations) as well as the base contents for all 4 nucleotides (A, C, G and U) as a function of time was used to infer the RNA-editing functions of ADAR and APOBEC proteins within the host. The enrichment of a specific type of mutations were tested using chisq tests.

### Mutation analysis related to the host response

The URL for data resources in investigating the nucleotide changes from Ruijin, Virginia, Spain, Wuhan and GISAID were detailed in Supplementary notes.

### Molecular Dynamics Simulation

The original structures (PDB format) of SARS-CoV-2 proteins were downloaded from Protein Data Bank (PDB, https://www.rcsb.org/) with accession numbers, ORF3a: 6xdc, Spike: 6vyb and NSP12:7bv2. Point mutations were introduced into each protein sequence and generated the mutated sequence. The mutated sequence and the corresponding original template protein structure were then taken as inputs for SWISS-MODEL for Homology modeling. After the modeling was completed, the PDB files of the target mutated proteins were obtained for further analysis. Subsequently, Ions and waters are deleted from PDB files. The PDB files were then subjected to GROMACS (Version: V5.1) and utilized for molecular dynamics simulation at the temperature 300K. Gromacs output the free energy (KJ/mol) to measure the stability of candidate protein. A smaller value of free energy indicates a higher stability of protein.

## Supporting information

MainTable1

## Data Availability

A total of 896 high quality consensus assemblies (with less than 2% gaps) were submitted to GISAID (EPI_ISL_698105-698169, EPI_ISL_698172-699161, EPI_ISL_708827-708838) and raw sequencing data aligned to the SARS-CoV-2 reference genome were uploaded to NCBI (PRJNA687136) . We combined our genomes with other publicly available sequences for a final dataset of 973 SARS-CoV-2 genomes(ncov_global.json, Supplementary file). The dataset can be visualized on the community Nextstrain page.

## Data availability

A total of 896 high quality consensus assemblies (with less than 2% gaps) were submitted to GISAID (EPI_ISL_698105-698169, EPI_ISL_698172-699161, EPI_ISL_708827-708838) and raw sequencing data aligned to the SARS-CoV-2 reference genome were uploaded to NCBI (PRJNA687136). We combined our genomes with other publicly available sequences for a final dataset of 973 SARS-CoV-2 genomes(ncov_global.json, Supplementary file). The dataset can be visualized on the ‘‘community’’ Nextstrain page.

## Acknowledgement

The study was supported by the Department of Health and SEHA in Abu Dhabi, the United Arab Emirates and National Natural Science Foundation of China (31900487). We would like to acknowledge Xihui Chen and Fengliang Cui, Qinglong Wang, Shengchang Gu, Guoyang Xu from MGI for the fast installation of sequencing instruments, the G42 Huoyan laboratory members who contributed to the collection of samples, Dr. Joseph Mafofo and Inacio Pacheco for suggestions on the IRB application, professors Huachun Zou, Qianglin Fang and associated professors Huicui Feng, Jinqiu Yuan and Yawen Jiang from the school of public health (Shenzhen), Sun-Yat-Sen University, associated professor Jinqiu Yuan from the seventh affiliated hospital and Dr. Min Sum Park for helpful scientific discussion and suggestion in the study.

## Author contributions

Conceptualization, S. Liu, W. Z; Methodology, J. L, S. Liu, R. Liu, P. W, K. L, P. L, L. L; Formal Analysis, R. Liu, P. W, D. L, W. H, S. Liu; Resources, S. M, T. M, Z. Y, X. M; Data Curation, R. Liu, N. K, M. F, H. K, J. Q, V. K; Writing - Original Fraft, S.Liu; Writing - Review & Editing, S.Liu, P. O, S. F, H. K, C.Y; Supervision, P. X, X. X, X. A, X. J, B. A, J. W, H. Y; Project Administration, T. M, F. C, N. Q, X. H and W. L; Funding Acquisition, A.K, W. L and S. Liu.

