## Supplementary material for "Genomic epidemiology of SARS-CoV-2 in the United Arab Emirates reveals novel virus mutation, patterns of co-infection and tissue specific host innate immune response": MainTable1

**Table 1. Allele frequency and functional annotation of the eleven UAE-specific genetic variants**

| **Position** | **UAE AF** | **Type** | **Region** | **Nucleotide change** | **Amino acid change** | **CNCB AF** | **P-value** |
| --- | --- | --- | --- | --- | --- | --- | --- |
| **5924** | 0.194 | missense | nsp3 | c.5659G>A | p.Val1887Ile | 0 | INF |
| **7171** | 0.091 | synonymous | nsp3 | c.6906T>C | p.Pro2302Pro | 0 | INF |
| **7851** | 0.069 | missense | nsp3 | c.7586C>T | p.Ala2529Val | 2.590E-04 | 3.943E-82 |
| **11230** | 0.203 | missense | nsp6 | c.10965G>T | p.Met3655Ile | 6.476E-04 | 3.108E-248 |
| **21775** | 0.206 | synonymous | S | c.213T>G | p.Ser71Ser | 0 | INF |
| **23311** | 0.089 | missense | S | c.1749G>T | p.Glu583Asp | 8.202E-04 | 1.250E-96 |
| **24170** | 0.065 | missense | S | c.2608A>G | p.Ile870Val | 4.317E-05 | 5.989E-84 |
| **27002** | 0.093 | synonymous | M | c.480C>T | p.Asp160Asp | 8.634E-05 | 1.058E-118 |
| **28167** | 0.182 | missense | ORF8 | c.274G>A | p.Glu92Lys | 4.317E-04 | 9.830E-226 |
| **28878** | 0.212 | missense | N | c.605G>A | p.Ser202Asn | 4.317E-04 | 2.31E-186 |
| **29742** | 0.235 | downstream | S | c.*4358G>A |  | 1.027E-02 | 1.658E-191 |

^#1^. Allele frequency computed from 896 genomes in UAE

^#2^. Allele frequency computed from 23,164 genomes around the globe

^#3^. Fisher exact test P-value comparing the allele counts between the 896 high quality UAE viral genomes and 23,164 viral genomes from the globe downloaded from the China National Center for Bioinformation. Comparison for all the 1,245 variants were detailed in Table S2.
