## SupplementaryInformation for "Genomic epidemiology of SARS-CoV-2 in the United Arab Emirates reveals novel virus mutation, patterns of co-infection and tissue specific host innate immune response"

### Supplementary Figures

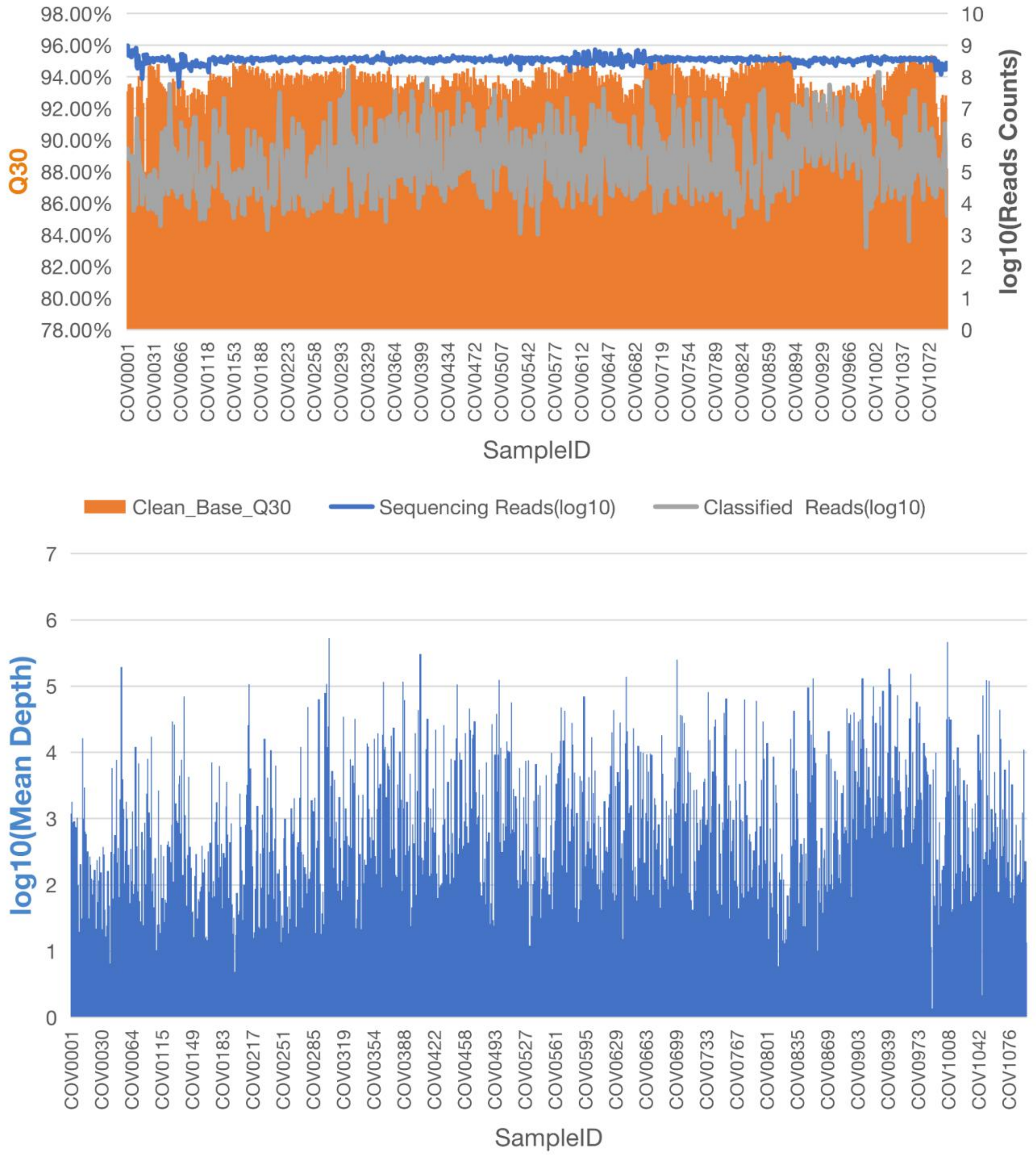

**Figure S1. Sequencing quality check.** (A) Percentage of sequencing bases reaching Q30 for each of the 1,067 samples is shown in orange bar. The base 10 logarithm of the number of sequencing reads and that of the number of reads classified as originated from SARS-CoV-2 is shown in blue and gray line, respectively. (B) The base 10 logarithm if the average sequencing depth of SARS-CoV-2 for each sample is shown in blue bar. The average sequencing depth ranges from 44 to 20,000 fold. X-axis shows the names of the 1,067 samples.

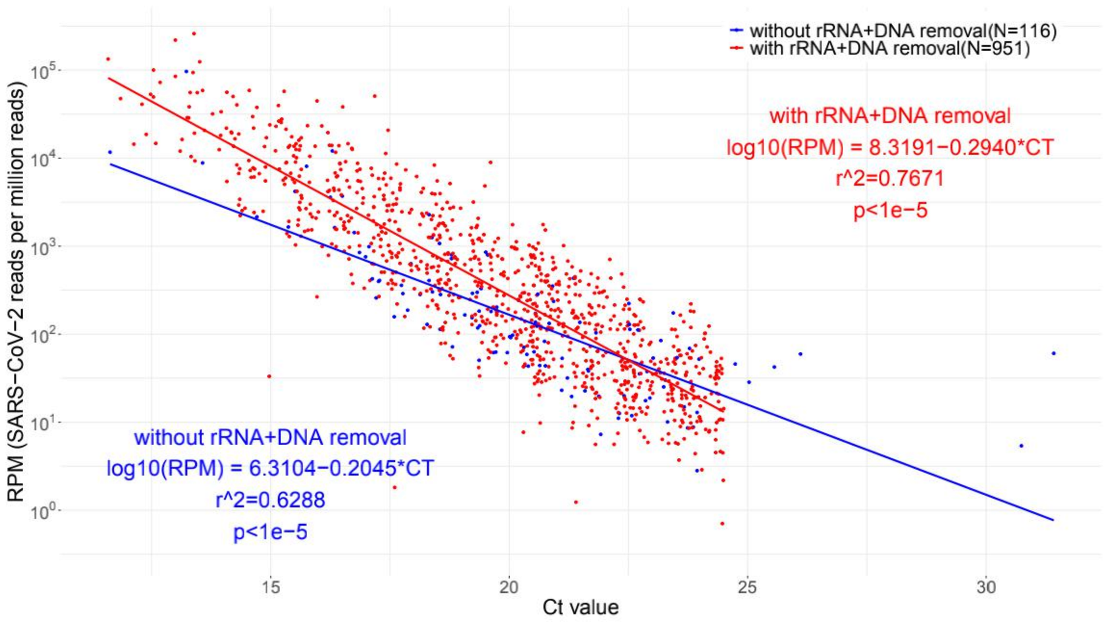

**Figure S2. Correlation between the relative viral load (RPM) estimated by meta-transcriptome sequencing and qRT-PCR Ct value using linear regression.** RPM indicates the number of reads aligned to SARS-CoV-2 reference per million total sequencing reads for each sample. There were two protocols adopted in the meta-transcriptomic sequencing. One without removal of rRNA and depletion of human DNA (N=116) while the other removes rRNA and deplete the human DNA during sequencing library prep (N=951). There is a high correlation between the Ct value and with the SARS-CoV-2 RPM estimated by both protocols ($R^2$~0.62-0.76). All the 1,067 positive samples have sequencing reads unambiguously aligned to the SARS-CoV-2 reference. All the 3 negative samples do have SARS-CoV-2 reads detected.

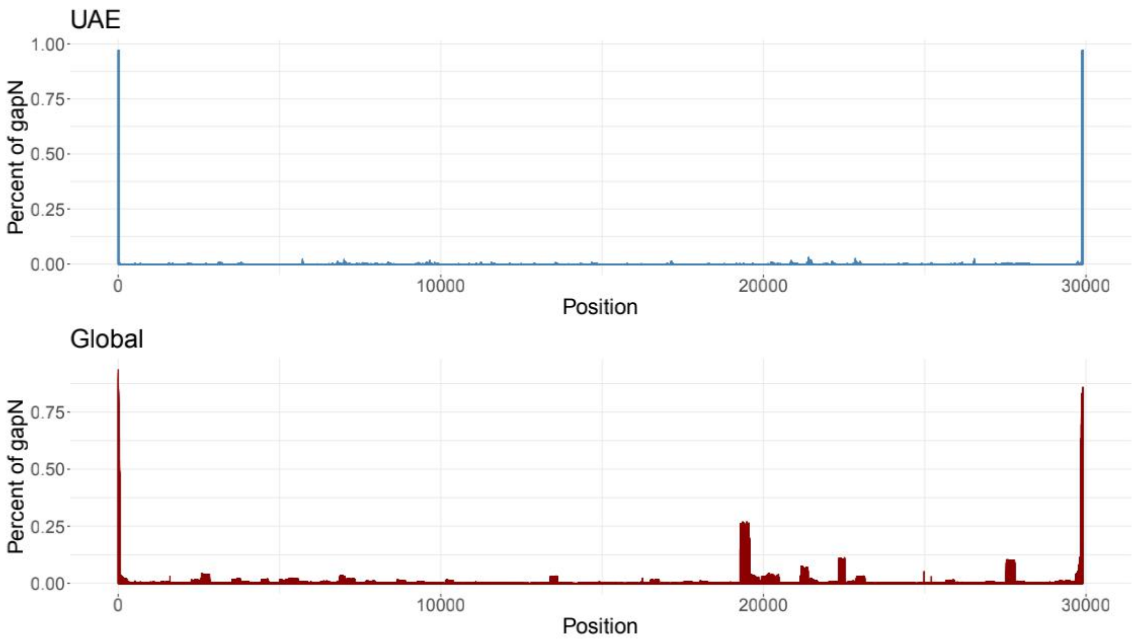

**Figure S3. Gap distribution over the SARS-CoV-2 reference genome.** (Top): gap distribution of the UAE SARS-CoV-2 assemblies reported in this study. (Bottom): gap distribution of the global assemblies reported in GISAID.

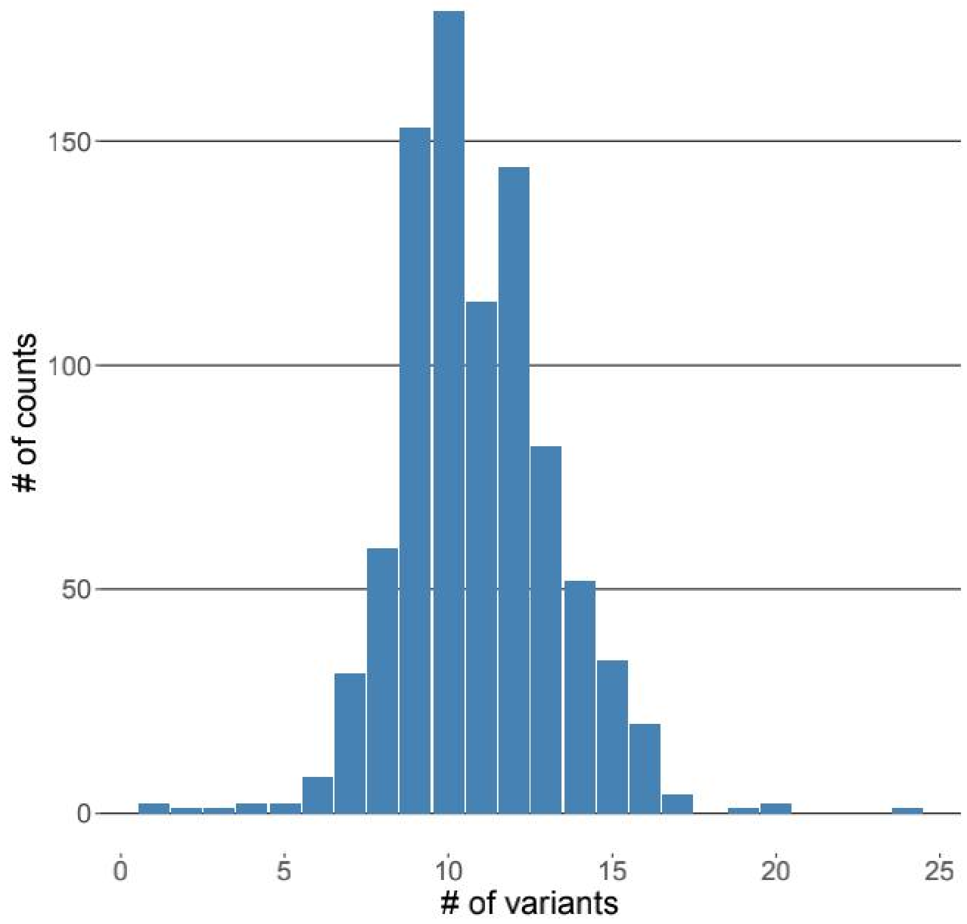

**Figure S4. Density distribution of the number of consensus variants per sample.** The variants were compared to the SARS-CoV-2 genome reference (BetaCoV/Wuhan/IVDC-HB-01/2019|EPI_ISL_402119).

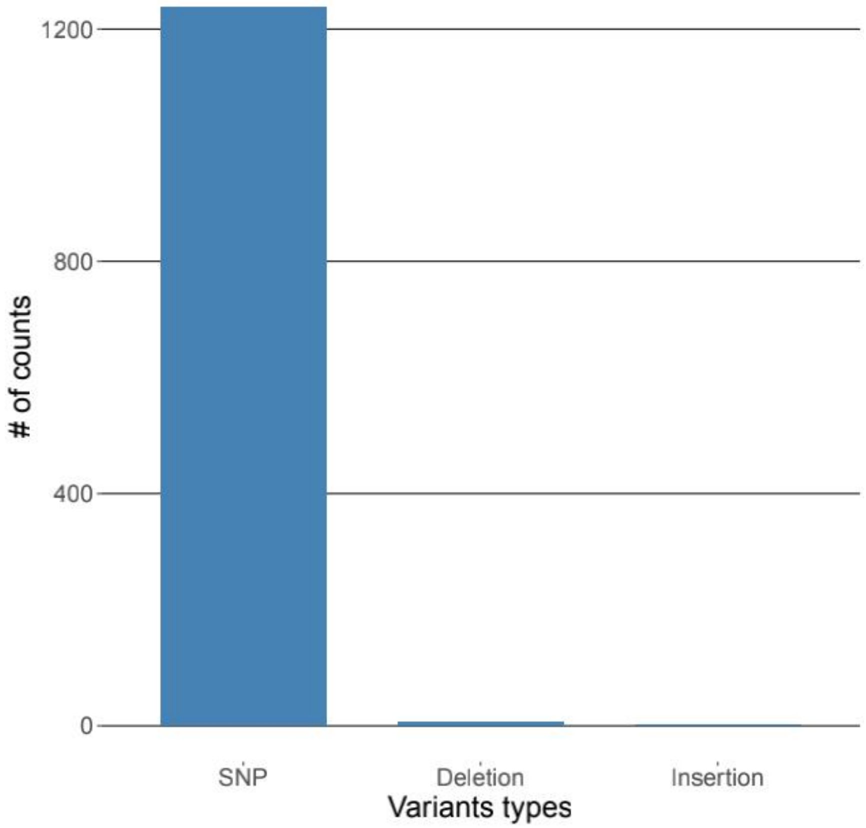

**Figure S5. Number of consensus variants identified from the 1,067 samples by variant type**. The consensus variants were detected by comparison to the SARS-CoV-2 genome reference (BetaCoV/Wuhan/IVDC-HB-01/2019|EPI_ISL_402119).

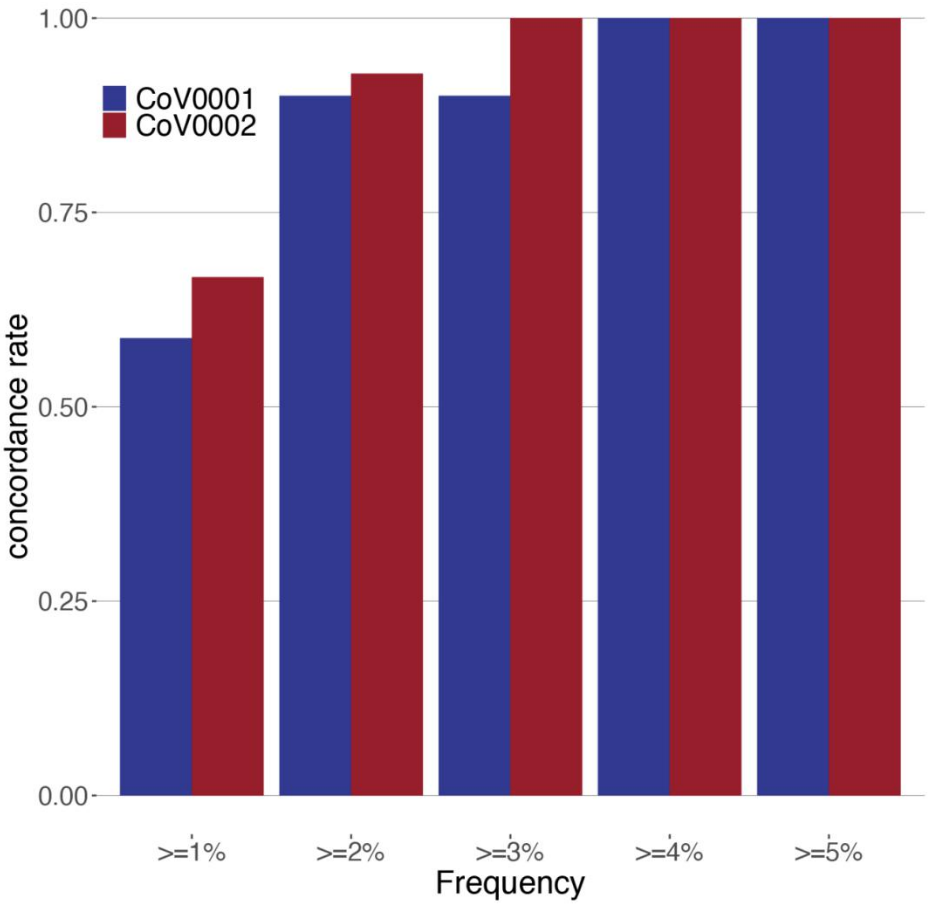

**Figure S6. Concordance rate of the intra-host variant detection as a function of increasing alternative allele frequency threshold.** Concordance rate was computed for the three technical replicates for the two randomly selected samples. Detailed statistics corresponding to this figure can be found in Table S2.

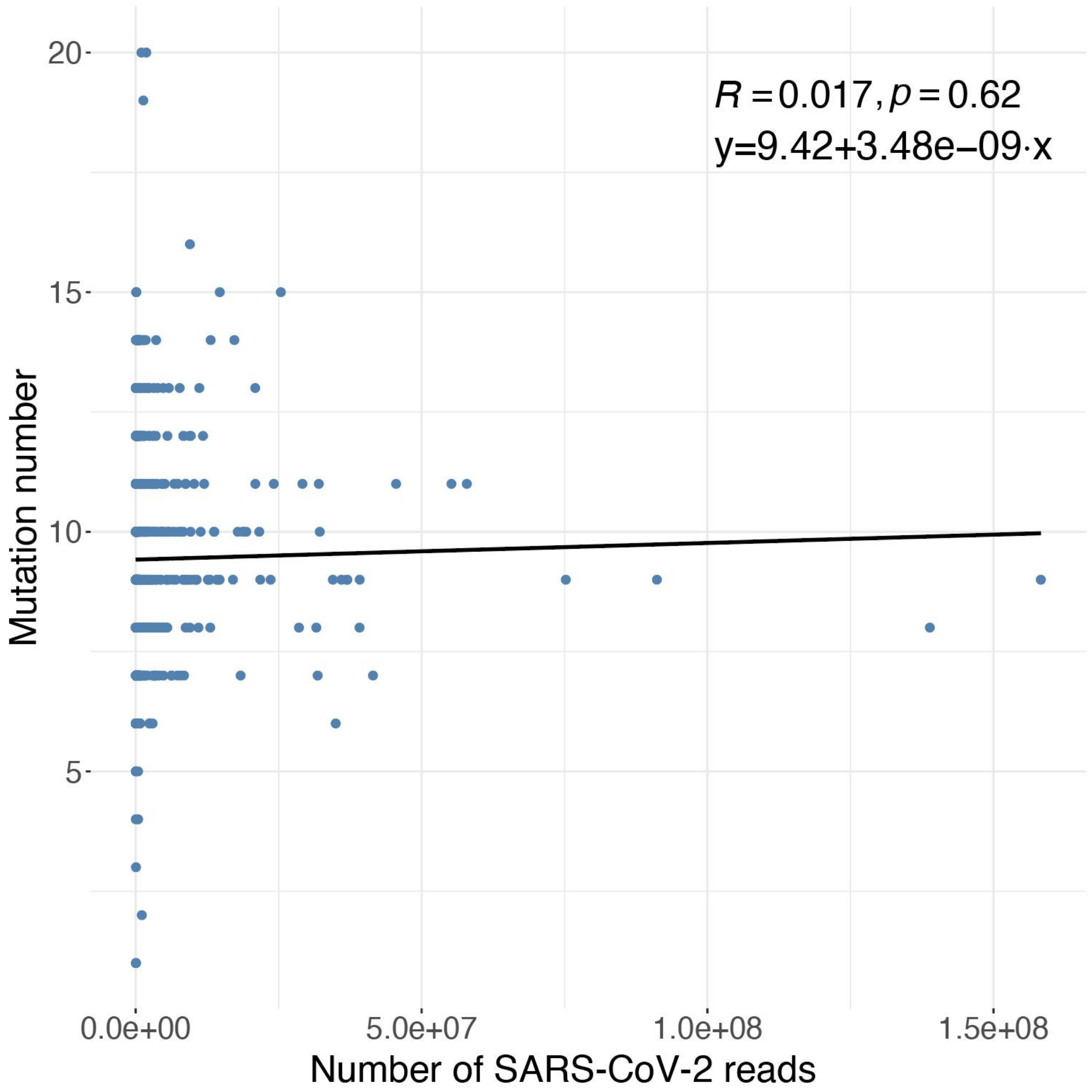

**Figure S7. Number of consensus variants identified per sample as a function of number of sequenced reads aligned to SARS-CoV-2.** The formula was derived from linear regression and the pearson correlation coefficient R was displayed.

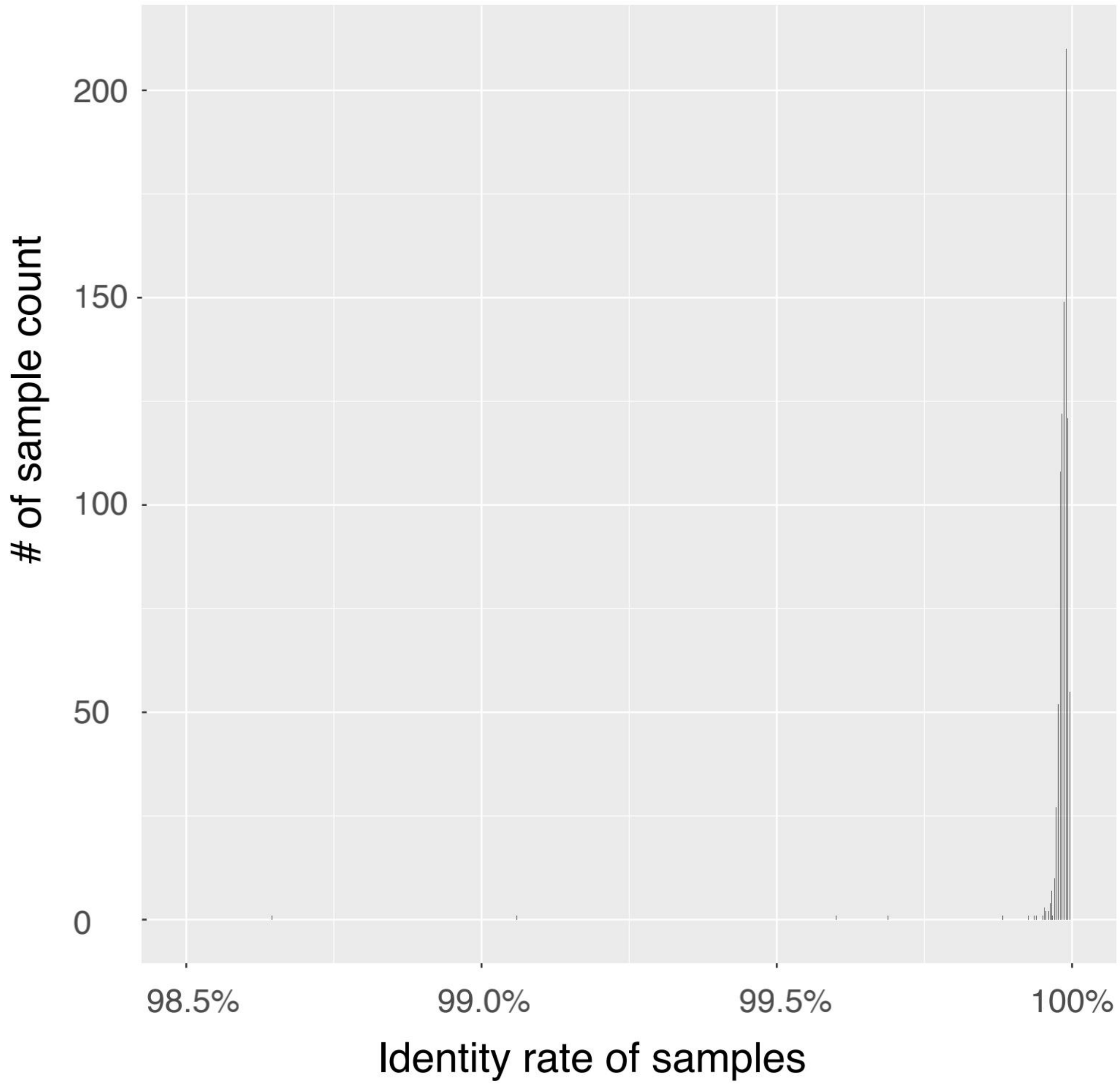

**Figure S8. Identity distribution of the 52 closest relative strains.** Shown is the density distribution of 52 closest relative strains compared with the 637 unique genomes assembled in our study.

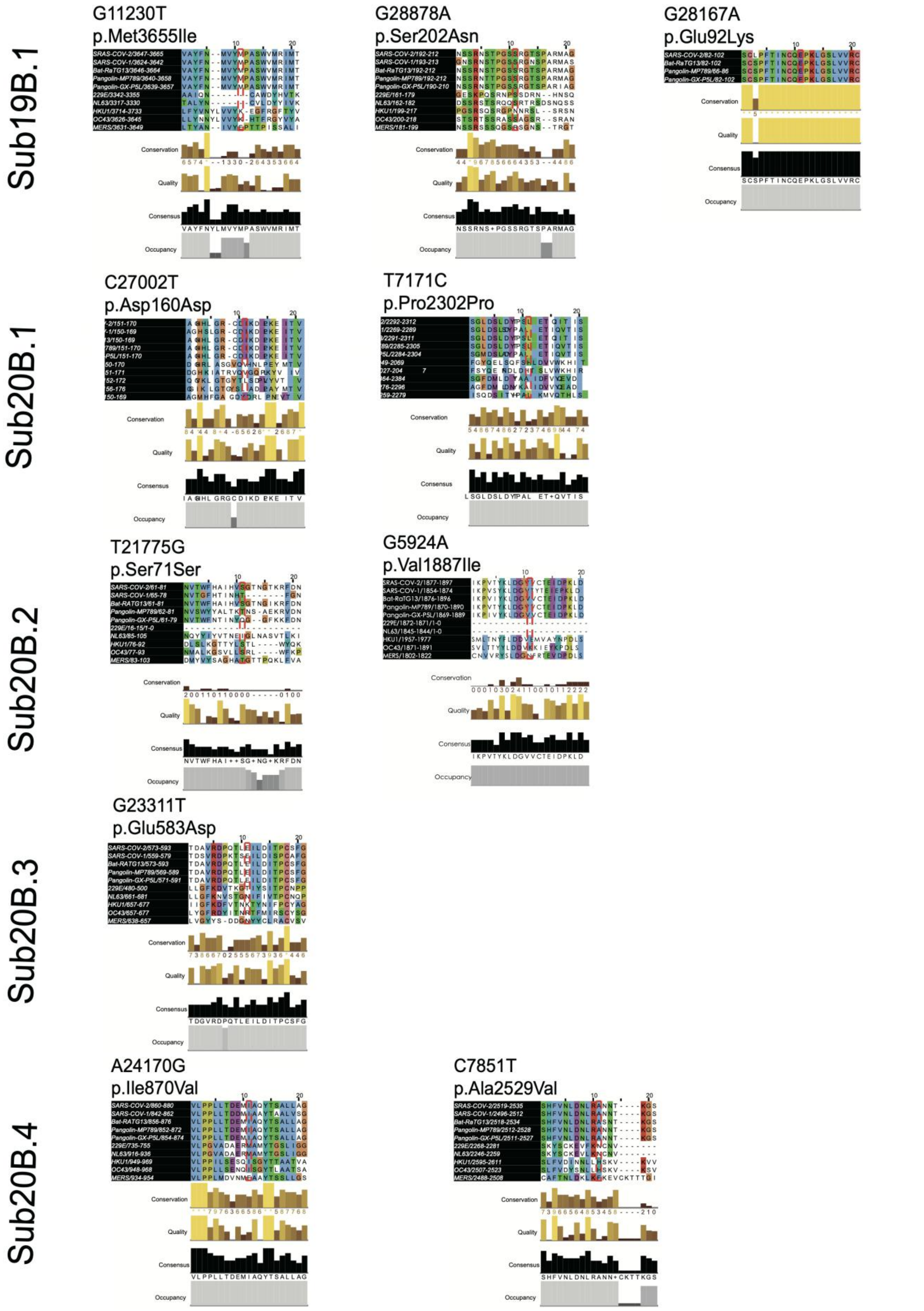

**Figure S9. Conservativeness of the eleven UAE sub-clade definitive genetic variation.** Shown is the conservation, quality, consensus and occupancy score for the amino acid sequence centering on the mutation. The calculation of the score was based on the multiple alignment of seven coronaviruses reference sequences using Jalview.

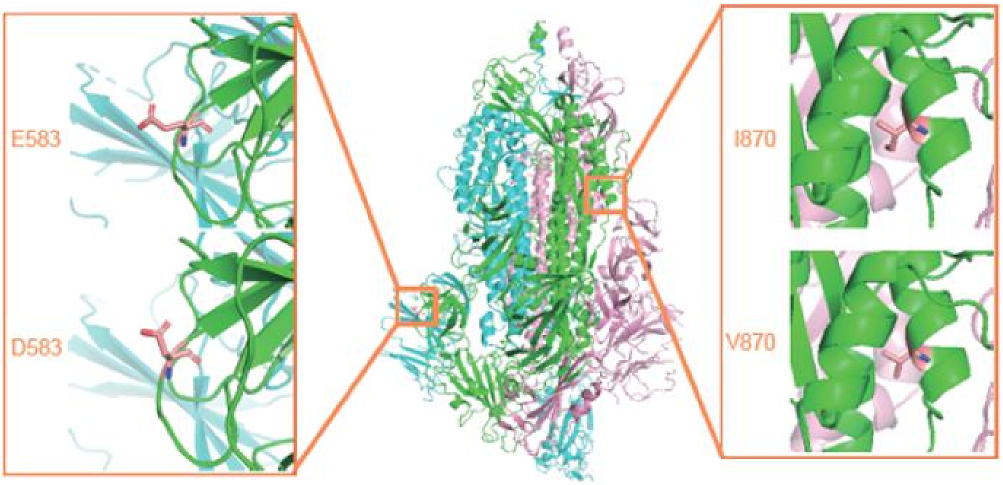

**Figure S10. Molecular dynamics analysis of the E583D and the I780V mutants in the S protein.** Shown are the changes of the protein structure comparing the wild types and the mutants.

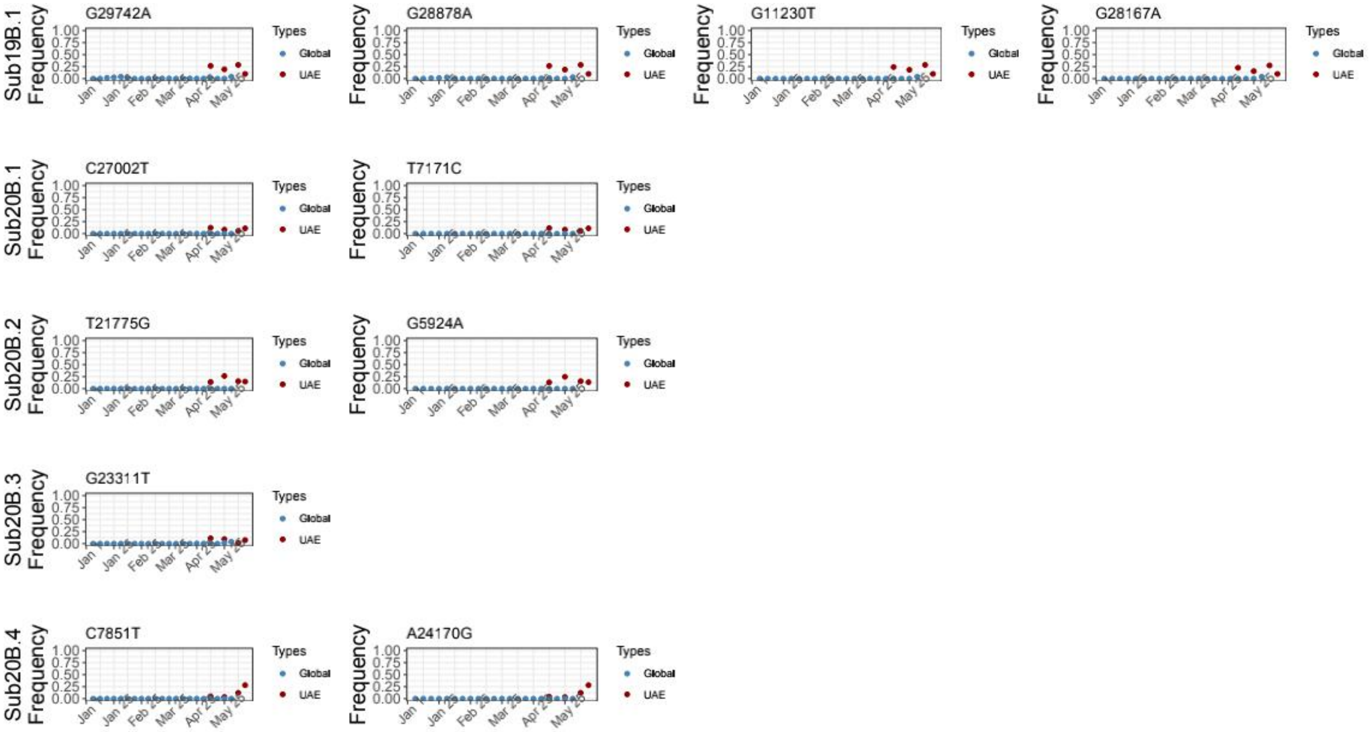

**Figure S11. Allele frequency changes of the subclade-definitive variants as a function of time**

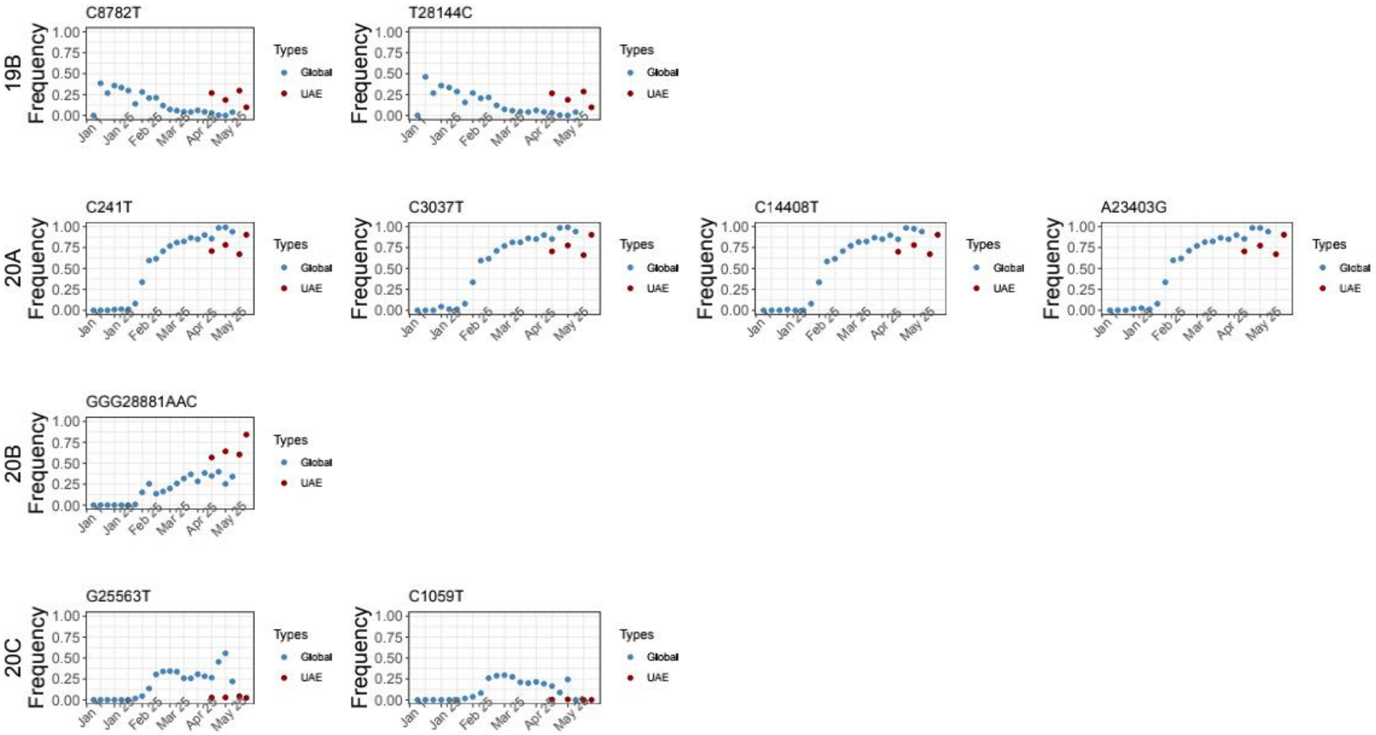

**Figure S12. Allele frequency changes of the clade-definitive variants as a function of time**

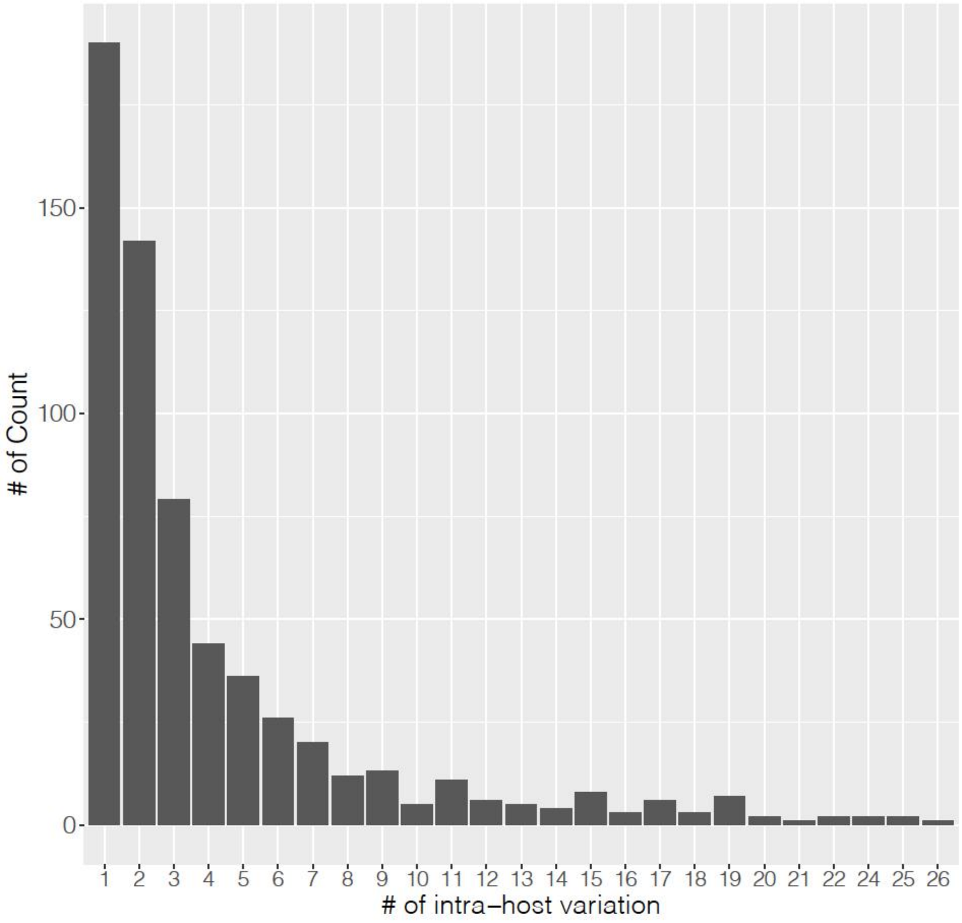

**Figure S13. Density distribution of number of intra-host variation per individual.** Number of iSNV with a 5% minor allele frequency threshold and a minimum of 4 minor allele counts was calculated for each individual.

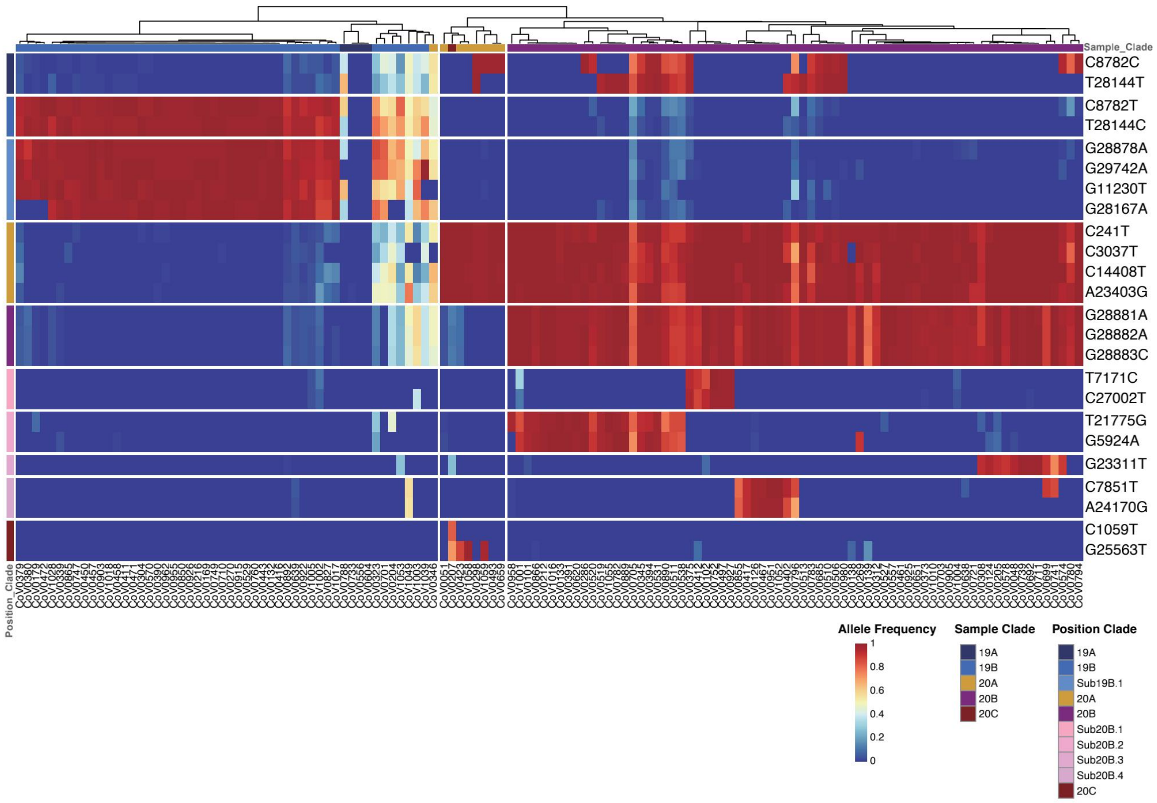

**Figure S14. Patterns of intra-host genetic variation.** The intra-host variation profile when using a 0.05% minor allele frequency threshold and a minimum of 4 minor allele counts per individual.

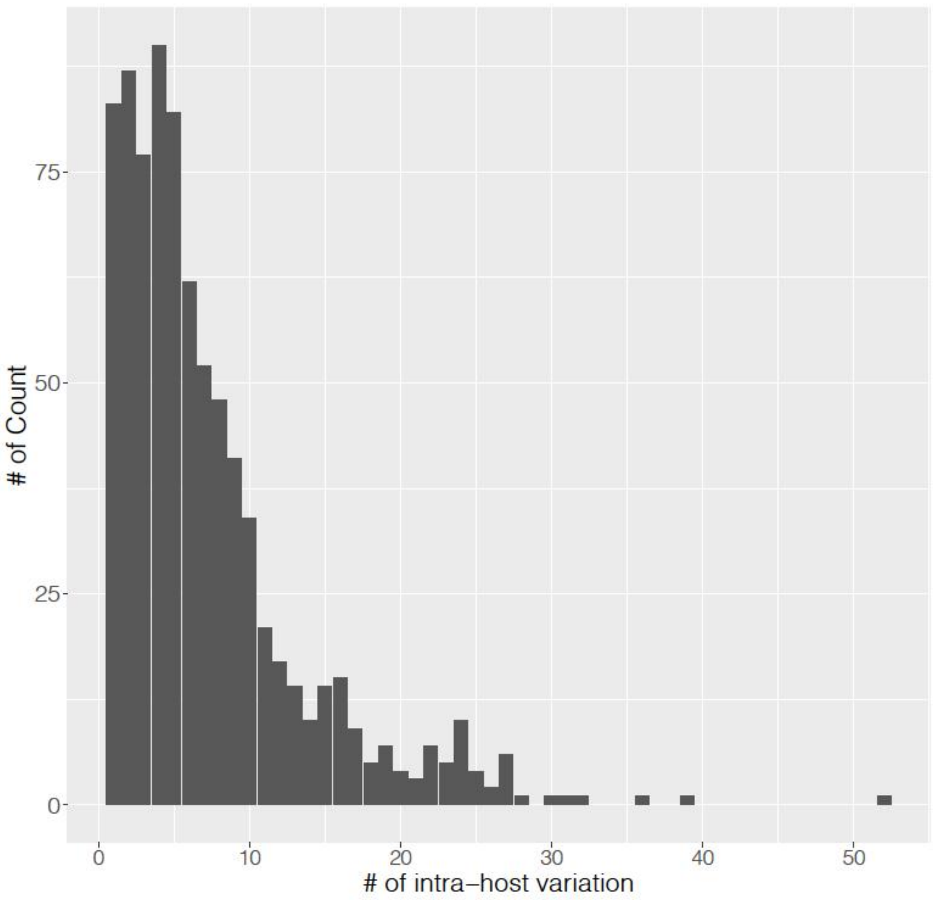

**Figure S15. Density distribution of number of intra-host variation per individual.** Number of iSNV with a 0.05% minor allele frequency threshold and a minimum of 4 minor allele counts was calculated for each individual.

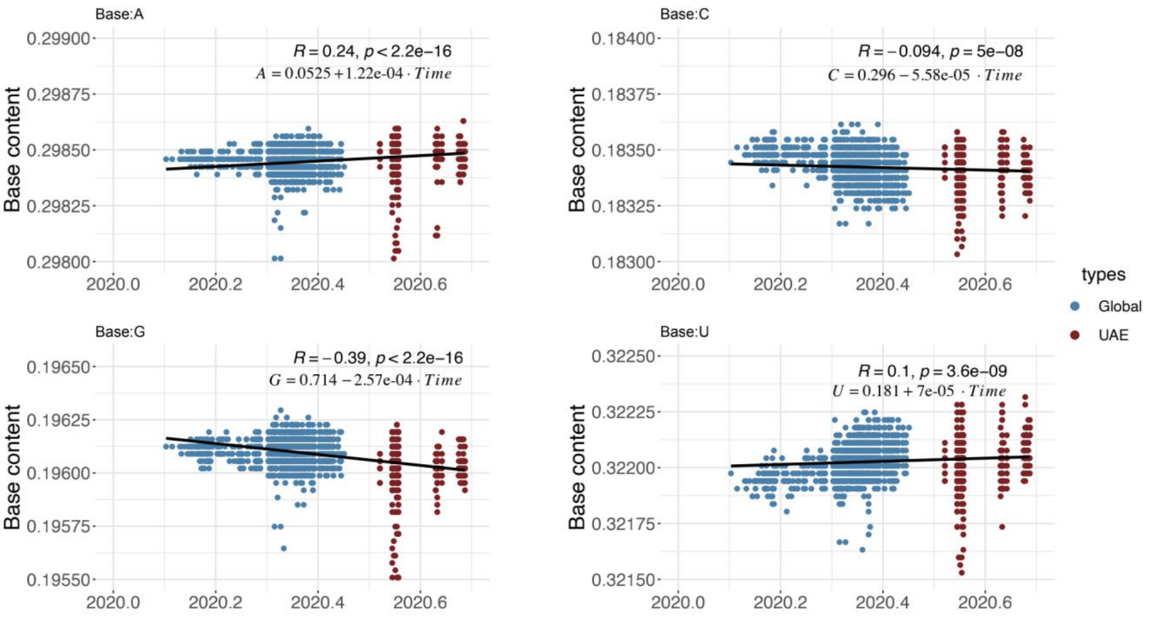

**Figure S16. Dynamic nucleotide composition at the coding regions of SARS-CoV-2 across time.** Each dot represents one sequence. The calculation was based on 2,574 unique SARS-CoV-2 strains isolated from December 24th, 2019 to April 17th, 2020 and the 896 high quality genomes in the UAE.

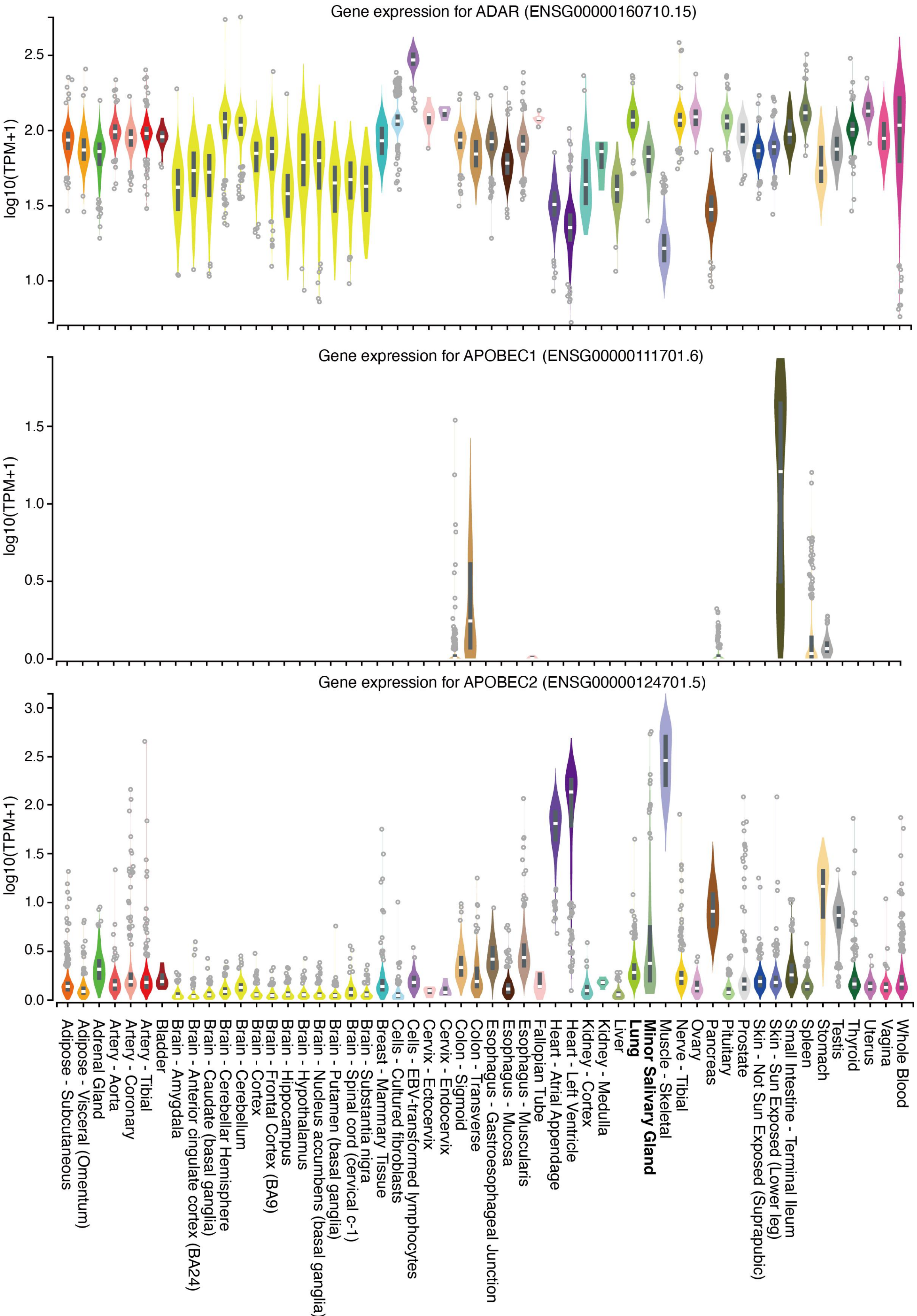

Gene expression for ADAR (ENSG00000160710.15)
log10(TPM+1)
2.5
2.0
1.5
1.0
Gene expression for APOBEC1 (ENSG00000111701.6)
log10(TPM+1)
1.5
1.0
0.5
0.0
Gene expression for APOBEC2 (ENSG00000124701.5)
log10(TPM+1)
3.0
2.5
2.0
1.5
1.0
0.5
0.0
Adipose - Subcutaneous
Adipose - Visceral (Omentum)
Adrenal Gland
Artery - Aorta
Artery - Coronary
Artery - Tibial
Bladder
Brain - Amygdala
Brain - Anterior cingulate cortex (BA24)
Brain - Caudate (basal ganglia)
Brain - Cerebellar Hemisphere
Brain - Cerebellum
Brain - Cortex
Brain - Frontal Cortex (BA9)
Brain - Hippocampus
Brain - Hypothalamus
Brain - Nucleus accumbens (basal ganglia)
Brain - Putamen (basal ganglia)
Brain - Spinal cord (cervical c-1)
Brain - Substantia nigra
Breast - Mammary Tissue
Cells - Cultured fibroblasts
Cells - EBV-transformed lymphocytes
Cervix - Ectocervix
Cervix - Endocervix
Colon - Sigmoid
Colon - Transverse
Esophagus - Gastroesophageal Junction
Esophagus - Mucosa
Esophagus - Muscularis
Fallopian Tube
Heart - Atrial Appendage
Heart - Left Ventricle
Kidney - Cortex
Kidney - Medulla
Liver
Lung
Minor Salivary Gland
Muscle - Skeletal
Nerve - Tibial
Ovary
Pancreas
Pituitary
Prostate
Skin - Not Sun Exposed (Suprapubic)
Skin - Sun Exposed (Lower leg)
Small Intestine - Terminal Ileum
Spleen
Stomach
Testis
Thyroid
Uterus
Vagina
Whole Blood

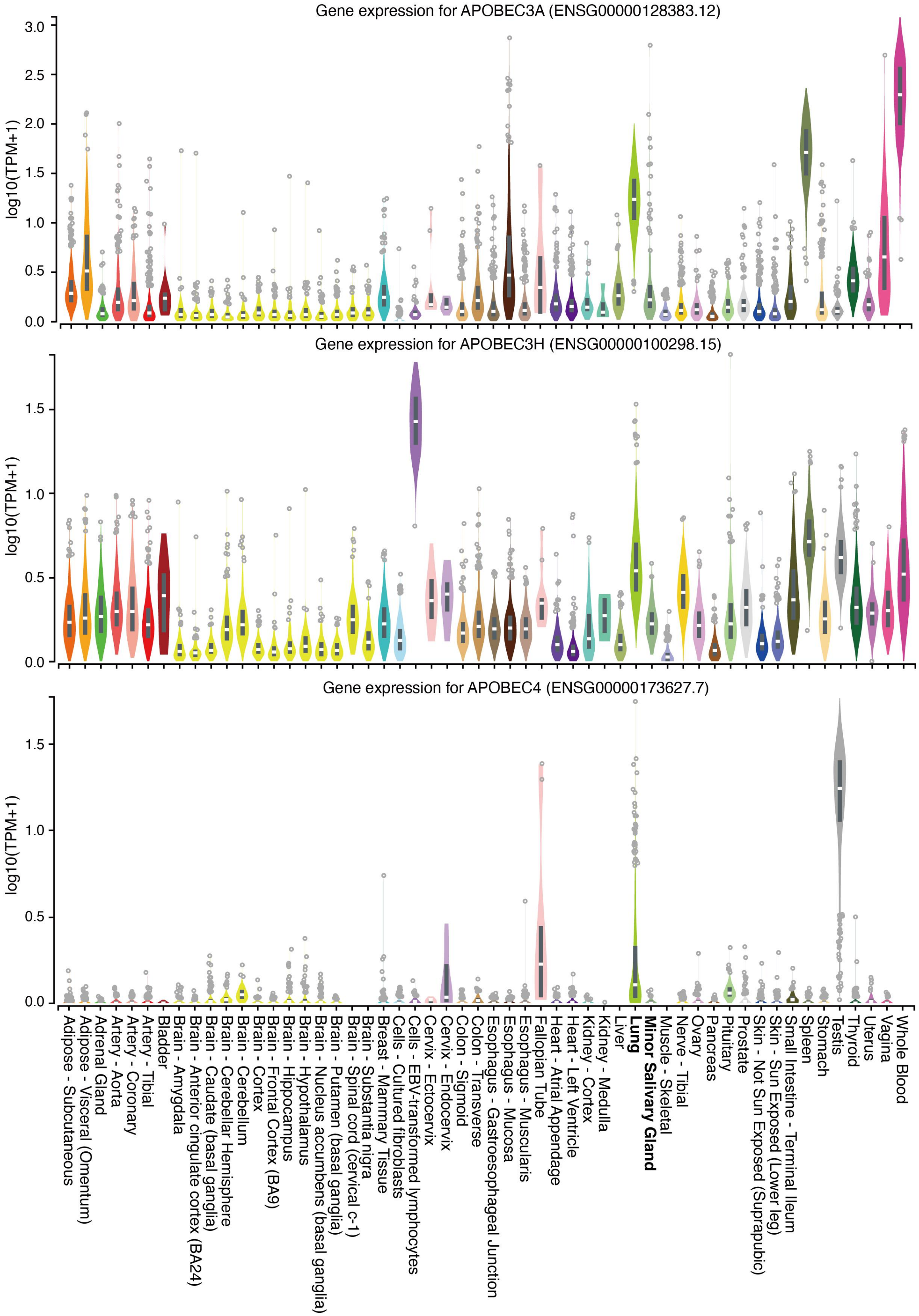

**Figure S17. Gene expression of ADAR gene among several issues from GTEX.**

### Supplementary Tables

**Table S1. Number of reads filtered during the analysis and coverage of the viral genome**

**Table S2. Information of the 1245 genetic variants identified from the 896 consensus viral assemblies**

**Table S3. Comparison of variants detected from Freebayes and Pilon and allele frequency estimated from REDITools between three technical replicates from two sample COV20R000001 and COV20R000002**

**Table S4. Samples that shared identical consensus genome sequence with the others**

**Table S5. Demographic data of 152 anonymous samples**

**Table S6. Closest relative in the GISAID database for the 896 high quality viral assemblies**

**Table S7. Molecular dynamic simutlation of the non-synonymous variants**

**Table S8. Wilcoxon test of gene expression difference**

### Supplementary Notes

#### Nomenclature of the clades and lineages

| Pangolin | GISAID | Clades |
| --- | --- | --- |
| A | S | 19B |
| B | L | 19A |
| B.2 | V | 19A |
| B.1 | G | 20A |
| B.1.* | GH | 20A/20C |
| B.1.1.1 | GR | 20B |
| B.1.177 | GV | 20A |

#### URL— Resource Table

| | **Select samples randomly** | **submission** | **URL** |
| --- | --- | --- | --- |
| **Ruijin** | 112/112 | Submitted to NCBI by Ruijin Hospital Affiliated to Shanghai Jiao Tong University School of Medicine | https://www.ncbi.nlm.nih.gov//bioproject/PRJNA627662 |
| **Virginia** | 35/474 | Submitted to NCBI by Virginia Division of Consolidated Laboratory Services Sequencing Submission Group | https://www.ncbi.nlm.nih.gov/bioproject/PRJNA625551 |
| **Spain** | 36/244 | Submitted to NCBI by FISABIO - Public Health | https://www.ncbi.nlm.nih.gov/bioproject/PRJEB37513 |
| **Wuhan** | 11/11 | Di Giorgio et al., Sci. Adv. 2020 | https://www.ncbi.nlm.nih.gov/bioproject/PRJNA601736<br>https://www.ncbi.nlm.nih.gov/bioproject/PRJNA603194<br>https://www.ncbi.nlm.nih.gov/bioproject/PRJNA605907 |
| **GISAID** | 23,164 | | https://bigd.big.ac.cn/ncov/variation/statistics |
